# Sociodemographic and health-related differences in non-diabetic hyperglycaemia and undiagnosed type 2 diabetes in the Health Survey for England 2013-2019

**DOI:** 10.1101/2024.02.12.24302519

**Authors:** Emma Campbell, Ellie Macey, Caitlin Kneebone-Hopkins, Chris Shine, Vahé Nafilyan, Nicholas Brown, Martin White, Ross Jones, Kamlesh Khunti, Katie Finning

**Affiliations:** Office for National Statistics, Government Buildings, Cardiff Rd, Duffryn, Newport NP10 8XG, UK; Department of Health and Social Care, London, UK; Diabetes UK, Wells Laurence House, 126 Back Church Lane, London, E1 1FH, UK; Diabetes Research Centre, University of Leicester, UK

**Keywords:** Diabetes, Prediabetes, Non-diabetic hyperglycaemia, Undiagnosed diabetes, Health inequalities

## Abstract

**Background:** Type 2 diabetes is a leading cause of morbidity and mortality but is often undiagnosed. Non-diabetic hyperglycaemia increases T2D risk, and its prevalence is increasing. Understanding the scale of undiagnosed T2D and NDH and identifying at-risk groups is important for detection and intervention.

**Methods:** We used the 2013-2019 Health Survey for England, an annual, nationally representative, cross-sectional survey. Respondents aged 16^+^ who were not pregnant and had valid HbA1c measurements were included. Outcomes were undiagnosed T2D (HbA1c ≥48 mmol/mol (6·5%) and no T2D diagnosis), and NDH (HbA1c 42-47 mmol/mol (6·0-6·4%) and no diagnosis). Age-adjusted prevalence was estimated across sociodemographic and health-related characteristics. Independent associations were assessed using logistic regression.

**Findings:** An estimated 2·25% (95% confidence interval 2·05-2·45) of adults had undiagnosed T2D and 11·54% (11·11-11·97) had NDH; an estimated 999,700 and 5,121,800 adults respectively. Prevalence was higher in those with known risk factors for T2D, such as older age or Black or Asian ethnicity. Among those with T2D, 30·21% (28·13-32·28) were undiagnosed. Younger adults and those self-reporting better health were most likely to be undiagnosed. For women, lower BMI, and waist circumference, not being prescribed antidepressants, and living in rural areas, were associated with increased likelihood of being undiagnosed.

**Interpretation:** Nearly a third of T2D cases were undiagnosed. Some groups less likely to have T2D were most likely to be undiagnosed. NDH is prevalent across all groups, even those considered low risk. Findings highlight the burden of T2D and NDH and emphasise the need for awareness across the population.

## Introduction

Rising prevalence of diabetes is a growing public health concern, driven largely by increases in type 2 diabetes (T2D), accounting for over 90% of cases.^1–3^ In 2019, T2D was estimated to affect 437·9 million individuals globally and was responsible for over 1·5 million deaths.^4^ In the UK, over 5 million people were estimated to have diabetes in 2021-22.^3^ Diabetes and related complications place a considerable burden on healthcare, costing the National Health Service (NHS) in England around £10 billion per year or 10% of the entire budget.^3^

T2D is associated with increased risk of microvascular complications and cardiovascular diseases such as strokes, heart attacks, and kidney disease.^1,5–7^ Early onset diabetes, which is associated with worse outcomes, is increasing in prevalence,^8^ and early detection and intervention are vitally important.^2,6,8^ However, T2D may be asymptomatic and an estimated 2-4% of adults in England had undiagnosed T2D between 2013-2019,^1,5,9^ with some evidence that undiagnosed T2D is associated with a higher cardiovascular risk than diagnosed diabetes.^10^ The proportion of T2D that is undiagnosed increased during the COVID-19 pandemic,^11^ with an estimated 60,000 missed or delayed diagnoses in the UK between March and December 2020.^12^ Non-diabetic hyperglycaemia (NDH), increases the risk of T2D and associated conditions,^13,14^ and refers to glycated haemoglobin (HbA1c) above normoglycaemic levels (<42 mmol/mol (6·0%)) but below the diabetes threshold (48 mmol/mol (6·5%)). NDH was estimated to affect 10·7% of adults in England between 2009-2013,^15^ but prevalence is increasing.^14^

Previous research found increased risk of diabetes in males, individuals belonging to South Asian and Black ethnic groups, older adults, those living in more deprived areas, and with health-related risk factors such as being overweight or having a high waist circumference.^1,2,9^ However, less is known about risk factors for undiagnosed diabetes. A previous study reported that older adults, those from non-white ethnic groups, in manual occupations, and with certain health markers such as higher cholesterol levels had an increased risk of undiagnosed diabetes.^16^ However, this study only included individuals aged 50^+^ and did not separate type 1 and T2D. Although previous research has demonstrated that individuals with NDH have similar demographic profiles to those with T2D,^13,14^ there is a lack of evidence on a wider range of NDH risk factors in the general population. Additionally, there is little evidence to establish whether risk factors for undiagnosed diabetes and NDH differ by sex, which is important given the known sex differences in T2D onset.^17^ This study aims to identify socioeconomic and health-related differences in the prevalence of NDH and undiagnosed T2D among a nationally representative sample of adults in England.

## Methods

### Study population

Data was pooled from the 2013-2019 Health Survey for England (HSE),^5^ an annual cross-sectional survey of those living in private households in England. HSE follows a multi-stage stratified probability sampling design involving a random selection of postcodes and addresses within postcodes. Sampling methods are available online.^5^ Participants completed an interview, including questions about health conditions, behaviours, and sociodemographic characteristics. Households may be selected for a nurse visit, where measurements and blood samples are taken. In 2013-2017 all participating households were offered nurse visits; from 2018 89% were randomly selected. Participants could decline the nurse visit and/or blood sampling.

Completion of the nurse visit from 2013-2019 ranged between 49-70%, and between 29-43% provided blood samples. Our final sample included 26,751 adults aged 16^+^, who were not pregnant, had valid HbA1c measurements, and answered the diabetes questions (see Supplementary Figure S1).

### Outcomes

#### Self-reported diagnosis of type 2 diabetes (T2D)

Participants were asked “Do you now have, or have you ever had diabetes?”. Those answering yes were asked “Were you told by a doctor that you had diabetes?” and “Have you been told by a doctor or nurse that you have type 1 or type 2 diabetes?”. We classified participants as having diagnosed T2D if they answered yes to the first two questions, confirmed a type 2 diagnosis, and that diabetes did not only occur during pregnancy. Some individuals (n = 93; 5% of those with a diabetes diagnosis) reported not knowing their diabetes type. Where these individuals’ age at diagnosis was 40^+^ (n=76), we classified them as having T2D since 90% of people diagnosed with diabetes have T2D, and type 1 is typically diagnosed earlier in life^18^. Where these individuals were diagnosed before the age of 40 (n=17) we categorised them as ‘type unknown’. Owing to differences in risk profiles and prevalence between type 1 diabetes and T2D, this analysis is restricted to T2D.

#### Glycated haemoglobin (HbA1c)

Participants without a previous diabetes diagnosis were categorised by their nurse visit HbA1c measurement. In September 2013 calibration changes impacted HbA1c results (full details in the HSE methods report).^5^. We used values adjusted to be comparable to pre-September 2013 for HbA1c measurements taken after recalibration, aligning with methods used by NHS Digital.

#### Undiagnosed T2D and non-diabetic hyperglycaemia (NDH)

We used self-reports of T2D and HbA1c measurements, in conjunction with National guidelines,^6^ to derive outcome groups:

1. Undiagnosed T2D – HbA1c 48 mmol/mol (6·5%) or above and no prior diabetes diagnosis; and
2. NDH – HbA1c between 42-47 mmol/mol (6·0-6·4%) with no prior diabetes diagnosis.

Undiagnosed T2D was explored both as prevalence among all adults, and as a percentage of those with T2D.

### Predictors

Predictors were informed by existing literature and data comparability and availability within the HSE. We included the following predictors: sex, age group, ethnicity, region, rural-urban classification, relationship status, highest educational qualification, National Statistics Socio-Economic Classification (NS-SEC), self-reported general health, antidepressant prescription, body mass index (BMI), waist circumference, smoking status, and alcohol consumption. Where age was the predictor, we used 10-year bands to ensure adequate sample sizes. An aggregated version was also created, which combined the 16-24-, 25-34-, and 35-44-year-old age groups, due to small numbers in these age groups with undiagnosed diabetes. Ethnicity was aggregated into ‘Black and Asian’ and ‘White, Mixed, and Other’ due to the known increased risk of T2D in Black and Asian ethnic groups, and to ensure adequate sample sizes. Data collection methods and variable levels for predictors are in Supplementary Table S1.

### Statistical analysis

All analyses accounted for the complex survey design and incorporated weights that account for selection, household and individual non-response, and refusal of nurse visit or blood sample (see HSE methods report for full weighting strategy).^5^ Weights were rescaled to the pooled dataset and sub-populations of interest by calculating the mean of the weight for each survey year within the sub-population and dividing the combined weights by their respective means.

For each predictor we estimated age-adjusted (a) prevalence of undiagnosed T2D, (b) proportion of adults with T2D who were undiagnosed, and (c) prevalence of NDH. Age adjustment involved fitting logistic regression models adjusted for age in five-year bands, the most granular level available, then using marginal means to estimate the age-adjusted prevalence within each predictor group. For each predictor-outcome combination we tested for moderation effects by sex, using interaction terms and Wald tests. Our main analysis was not stratified by sex due to sample size limitations, but interaction tests and stratified results are presented in Supplementary Material.

We examined the odds of (a) T2D being undiagnosed (versus diagnosed, among adults with T2D) and (b) having NDH (versus not having NDH, among adults without diabetes), after adjusting for other factors. For each predictor we fitted logistic regression models adjusted for age, sex, ethnicity, relationship status, rural-urban classification, region, and highest educational qualification. Confounders were selected based on previous research and topic knowledge, excluding variables likely to be on the causal pathway between the predictors and outcomes, and those likely to be collinear.

Population estimates and 95% confidence intervals for the number of people with undiagnosed T2D and NDH were calculated by applying weighted estimated proportions to the ONS mid-year population estimate for 2016 (the mid-year of our sample), adjusted to represent adults living in private households in England. This method is recommended in HSE methods report.^5^ Population estimates are rounded to the nearest 100.

There were negligible missing data (<0·2%) for all variables (Supplementary Table S2), except BMI (8·6%), therefore analyses were performed on a complete case basis. BMI was not included as a covariate in adjusted models therefore the missing data only impacted analysis where BMI was the predictor.

Data were analysed in R V4·1·3.

### Ethics

The UK Statistics Authority Data Ethics Team granted approval for this analysis. Respondents provided verbal informed consent for the interview and written consent for biological samples.

## Results

The sample included 26,751 adults in England, 54·86% were female and 7·58% were from a Black or Asian ethnic group (Table 1). An estimated 7·44% (95% confidence interval 7·07-7·82) of adults had type 2 diabetes (T2D), with higher prevalence among men (8·51% (7·93-9·09)) than women (6·42% (6·00-6·84)). Interaction tests and sex-stratified estimates are in Supplementary Tables S3-S6 and Figures S2-S5. We focus on non-stratified results but reference stratified results where sex differences were observed.

**Table 1.**
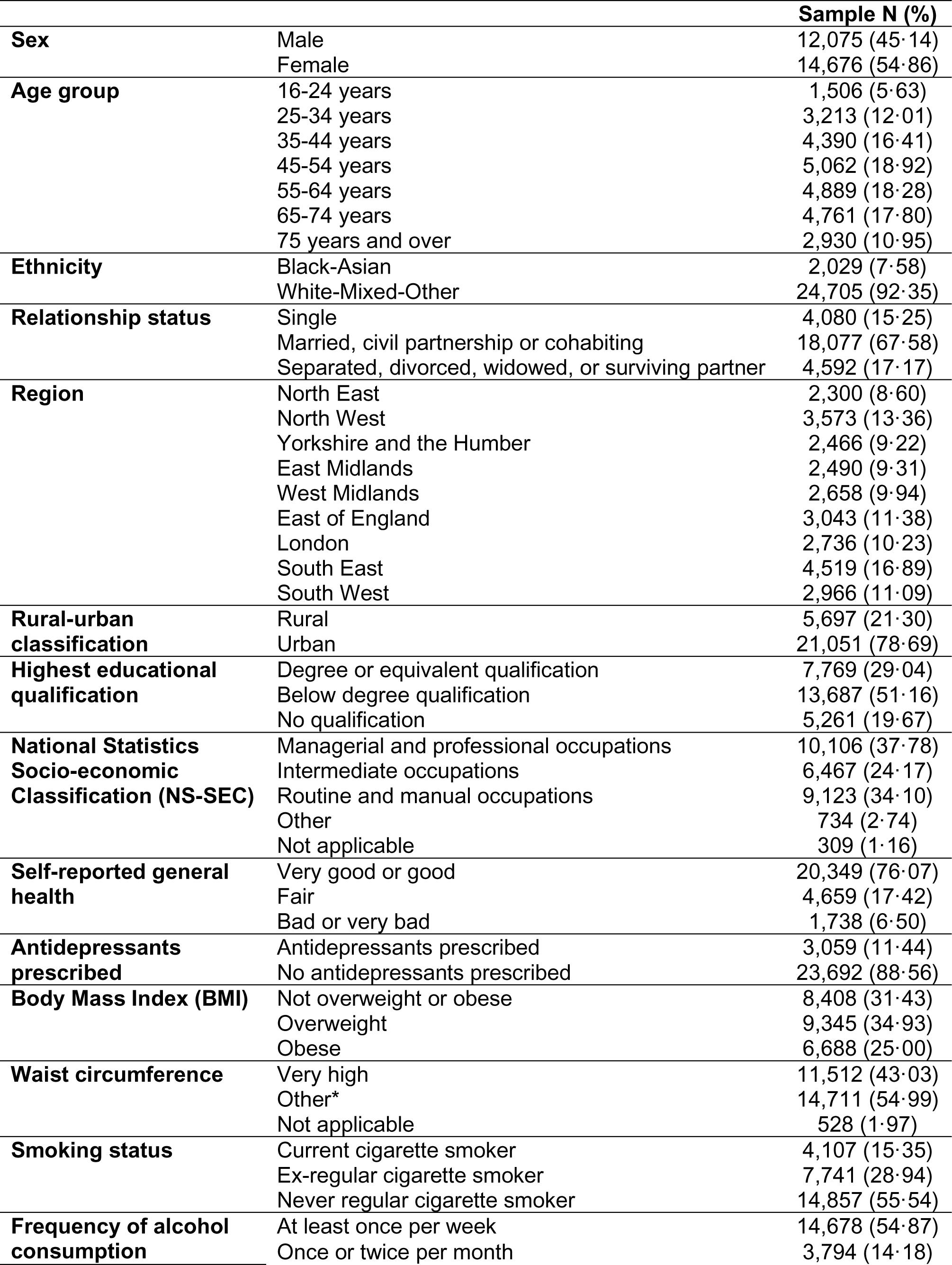
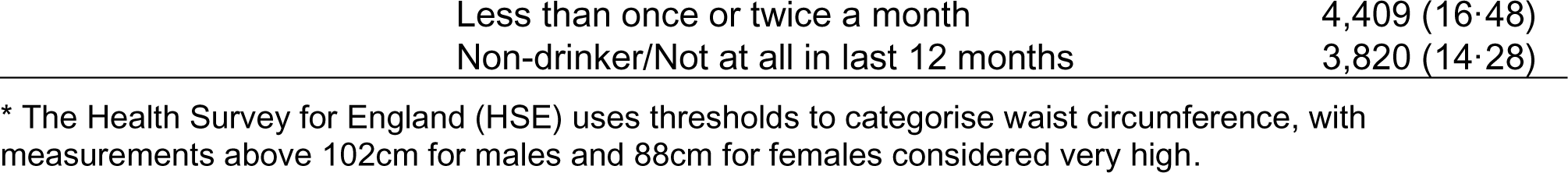
Characteristics of the sample.

|  |  | **Sample N (%)** |
| --- | --- | --- |
| **Sex** | Male | 12,075 (45.14) |
|  | Female | 14,676 (54.86) |
| **Age group** | 16-24 years | 1,506 (5.63) |
|  | 25-34 years | 3,213 (12.01) |
|  | 35-44 years | 4,390 (16.41) |
|  | 45-54 years | 5,062 (18.92) |
|  | 55-64 years | 4,889 (18.28) |
|  | 65-74 years | 4,761 (17.80) |
|  | 75 years and over | 2,930 (10.95) |
| **Ethnicity** | Black-Asian | 2,029 (7.58) |
|  | White-Mixed-Other | 24,705 (92.35) |
| **Relationship status** | Single | 4,080 (15.25) |
|  | Married, civil partnership or cohabiting | 18,077 (67.58) |
|  | Separated, divorced, widowed, or surviving partner | 4,592 (17.17) |
| **Region** | North East | 2,300 (8.60) |
|  | North West | 3,573 (13.36) |
|  | Yorkshire and the Humber | 2,466 (9.22) |
|  | East Midlands | 2,490 (9.31) |
|  | West Midlands | 2,658 (9.94) |
|  | East of England | 3,043 (11.38) |
|  | London | 2,736 (10.23) |
|  | South East | 4,519 (16.89) |
|  | South West | 2,966 (11.09) |
| **Rural-urban classification** | Rural | 5,697 (21.30) |
|  | Urban | 21,051 (78.69) |
| **Highest educational qualification** | Degree or equivalent qualification | 7,769 (29.04) |
|  | Below degree qualification | 13,687 (51.16) |
|  | No qualification | 5,261 (19.67) |
| **National Statistics Socio-economic Classification (NS-SEC)** | Managerial and professional occupations | 10,106 (37.78) |
|  | Intermediate occupations | 6,467 (24.17) |
|  | Routine and manual occupations | 9,123 (34.10) |
|  | Other | 734 (2.74) |
|  | Not applicable | 309 (1.16) |
| **Self-reported general health** | Very good or good | 20,349 (76.07) |
|  | Fair | 4,659 (17.42) |
|  | Bad or very bad | 1,738 (6.50) |
| **Antidepressants prescribed** | Antidepressants prescribed | 3,059 (11.44) |
|  | No antidepressants prescribed | 23,692 (88.56) |
| **Body Mass Index (BMI)** | Not overweight or obese | 8,408 (31.43) |
|  | Overweight | 9,345 (34.93) |
|  | Obese | 6,688 (25.00) |
| **Waist circumference** | Very high | 11,512 (43.03) |
|  | Other* | 14,711 (54.99) |
|  | Not applicable | 528 (1.97) |
| **Smoking status** | Current cigarette smoker | 4,107 (15.35) |
|  | Ex-regular cigarette smoker | 7,741 (28.94) |
|  | Never regular cigarette smoker | 14,857 (55.54) |
| **Frequency of alcohol consumption** | At least once per week | 14,678 (54.87) |
|  | Once or twice per month | 3,794 (14.18) |

|  |  |
| --- | --- |
| Less than once or twice a month | 4,409 (16.48) |
| Non-drinker/Not at all in last 12 months | 3,820 (14.28) |

\* The Health Survey for England (HSE) uses thresholds to categorise waist circumference, with measurements above 102cm for males and 88cm for females considered very high.

### Undiagnosed T2D among all adults

An estimated 2·25% (2·05-2·45) of adults had undiagnosed T2D, equating to 999,700 (910,200-1,089,200) individuals living in private households in England. There was a higher prevalence among men (2·57% (2·25-2·90) versus 1·94% (1·72-2·17) in women) and older adults (5·54% (4·64-6·45) aged 75 years and above versus 0·70% (0·48-0·92) aged 16-44) (Table 2).

**Table 2.**
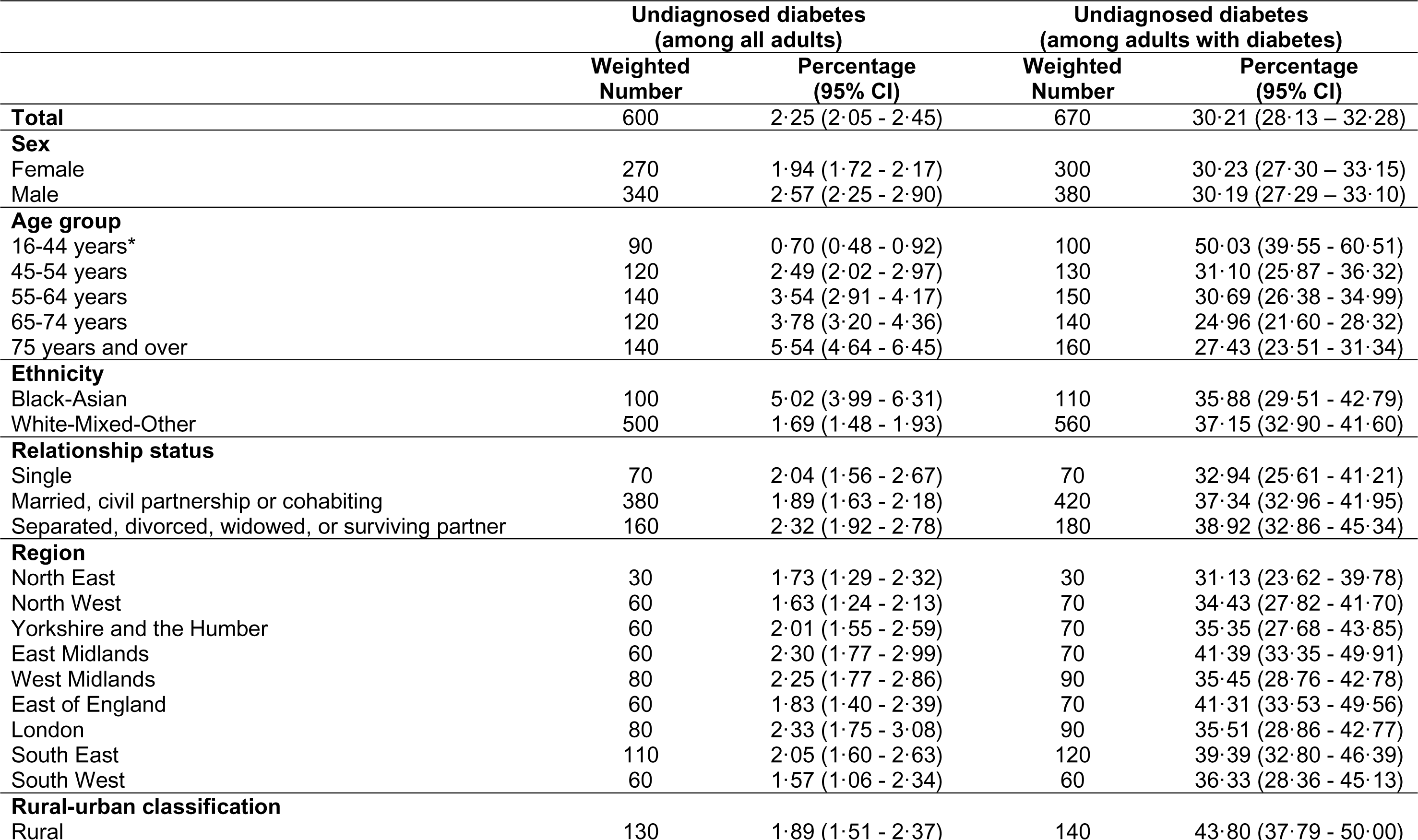
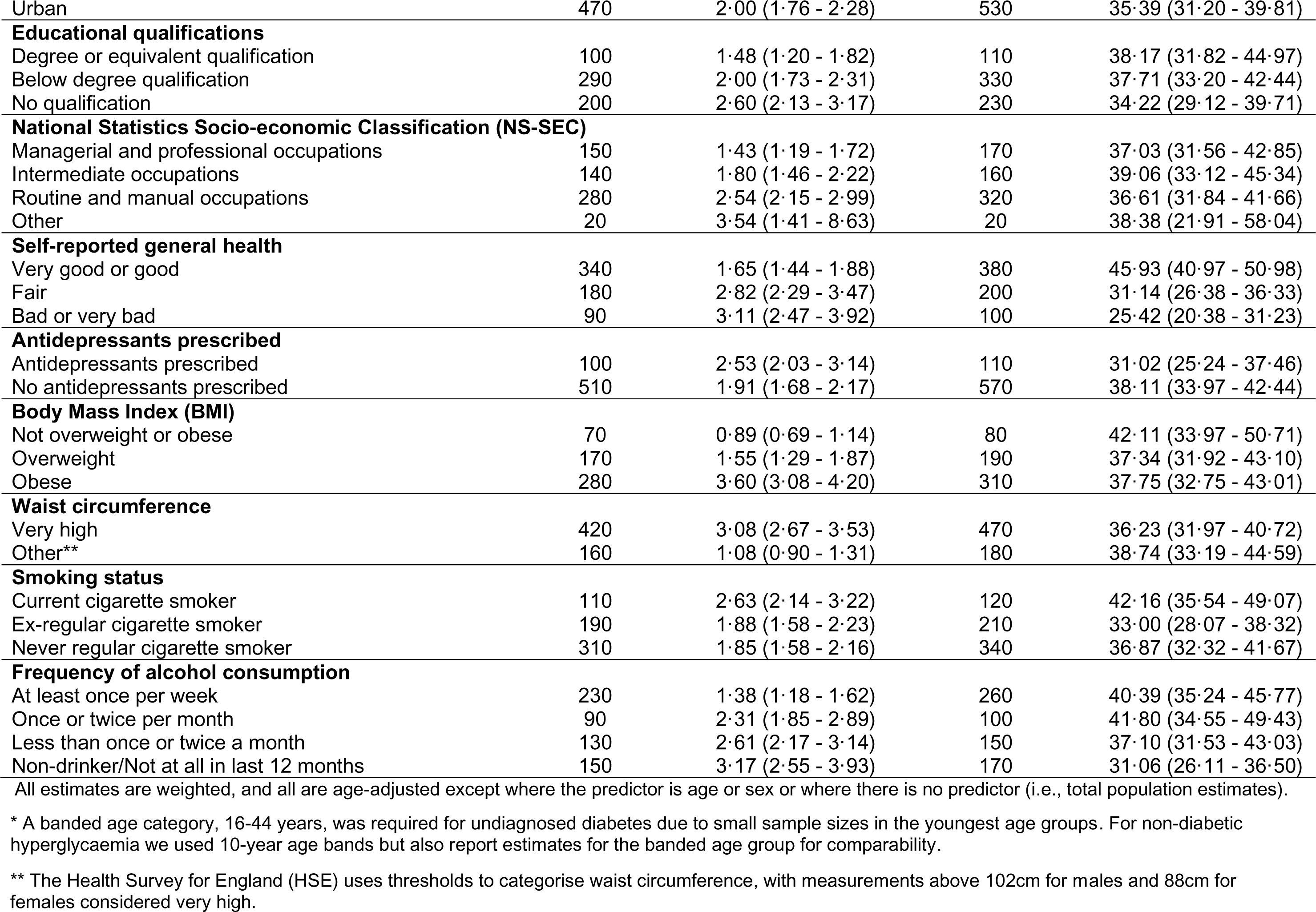
Percentage with undiagnosed type 2 diabetes by characteristics.

|  | Undiagnosed diabetes<br>(among all adults) |  | Undiagnosed diabetes<br>(among adults with diabetes) |  |
| --- | --- | --- | --- | --- |
|  | Weighted<br>Number | Percentage<br>(95% CI) | Weighted<br>Number | Percentage<br>(95% CI) |
| **Total** | 600 | 2.25 (2.05 - 2.45) | 670 | 30.21 (28.13 – 32.28) |
| **Sex** |  |  |  |  |
| Female | 270 | 1.94 (1.72 - 2.17) | 300 | 30.23 (27.30 – 33.15) |
| Male | 340 | 2.57 (2.25 - 2.90) | 380 | 30.19 (27.29 – 33.10) |
| **Age group** |  |  |  |  |
| 16-44 years* | 90 | 0.70 (0.48 - 0.92) | 100 | 50.03 (39.55 - 60.51) |
| 45-54 years | 120 | 2.49 (2.02 - 2.97) | 130 | 31.10 (25.87 - 36.32) |
| 55-64 years | 140 | 3.54 (2.91 - 4.17) | 150 | 30.69 (26.38 - 34.99) |
| 65-74 years | 120 | 3.78 (3.20 - 4.36) | 140 | 24.96 (21.60 - 28.32) |
| 75 years and over | 140 | 5.54 (4.64 - 6.45) | 160 | 27.43 (23.51 - 31.34) |
| **Ethnicity** |  |  |  |  |
| Black-Asian | 100 | 5.02 (3.99 - 6.31) | 110 | 35.88 (29.51 - 42.79) |
| White-Mixed-Other | 500 | 1.69 (1.48 - 1.93) | 560 | 37.15 (32.90 - 41.60) |
| **Relationship status** |  |  |  |  |
| Single | 70 | 2.04 (1.56 - 2.67) | 70 | 32.94 (25.61 - 41.21) |
| Married, civil partnership or cohabiting | 380 | 1.89 (1.63 - 2.18) | 420 | 37.34 (32.96 - 41.95) |
| Separated, divorced, widowed, or surviving partner | 160 | 2.32 (1.92 - 2.78) | 180 | 38.92 (32.86 - 45.34) |
| **Region** |  |  |  |  |
| North East | 30 | 1.73 (1.29 - 2.32) | 30 | 31.13 (23.62 - 39.78) |
| North West | 60 | 1.63 (1.24 - 2.13) | 70 | 34.43 (27.82 - 41.70) |
| Yorkshire and the Humber | 60 | 2.01 (1.55 - 2.59) | 70 | 35.35 (27.68 - 43.85) |
| East Midlands | 60 | 2.30 (1.77 - 2.99) | 70 | 41.39 (33.35 - 49.91) |
| West Midlands | 80 | 2.25 (1.77 - 2.86) | 90 | 35.45 (28.76 - 42.78) |
| East of England | 60 | 1.83 (1.40 - 2.39) | 70 | 41.31 (33.53 - 49.56) |
| London | 80 | 2.33 (1.75 - 3.08) | 90 | 35.51 (28.86 - 42.77) |
| South East | 110 | 2.05 (1.60 - 2.63) | 120 | 39.39 (32.80 - 46.39) |
| South West | 60 | 1.57 (1.06 - 2.34) | 60 | 36.33 (28.36 - 45.13) |
| **Rural-urban classification** |  |  |  |  |
| Rural | 130 | 1.89 (1.51 - 2.37) | 140 | 43.80 (37.79 - 50.00) |
| Urban | 470 | 2.00 (1.76 - 2.28) | 530 | 35.39 (31.20 - 39.81) |
| **Educational qualifications** |  |  |  |  |
| Degree or equivalent qualification | 100 | 1.48 (1.20 - 1.82) | 110 | 38.17 (31.82 - 44.97) |
| Below degree qualification | 290 | 2.00 (1.73 - 2.31) | 330 | 37.71 (33.20 - 42.44) |
| No qualification | 200 | 2.60 (2.13 - 3.17) | 230 | 34.22 (29.12 - 39.71) |
| **National Statistics Socio-economic Classification (NS-SEC)** |  |  |  |  |
| Managerial and professional occupations | 150 | 1.43 (1.19 - 1.72) | 170 | 37.03 (31.56 - 42.85) |
| Intermediate occupations | 140 | 1.80 (1.46 - 2.22) | 160 | 39.06 (33.12 - 45.34) |
| Routine and manual occupations | 280 | 2.54 (2.15 - 2.99) | 320 | 36.61 (31.84 - 41.66) |
| Other | 20 | 3.54 (1.41 - 8.63) | 20 | 38.38 (21.91 - 58.04) |
| **Self-reported general health** |  |  |  |  |
| Very good or good | 340 | 1.65 (1.44 - 1.88) | 380 | 45.93 (40.97 - 50.98) |
| Fair | 180 | 2.82 (2.29 - 3.47) | 200 | 31.14 (26.38 - 36.33) |
| Bad or very bad | 90 | 3.11 (2.47 - 3.92) | 100 | 25.42 (20.38 - 31.23) |
| **Antidepressants prescribed** |  |  |  |  |
| Antidepressants prescribed | 100 | 2.53 (2.03 - 3.14) | 110 | 31.02 (25.24 - 37.46) |
| No antidepressants prescribed | 510 | 1.91 (1.68 - 2.17) | 570 | 38.11 (33.97 - 42.44) |
| **Body Mass Index (BMI)** |  |  |  |  |
| Not overweight or obese | 70 | 0.89 (0.69 - 1.14) | 80 | 42.11 (33.97 - 50.71) |
| Overweight | 170 | 1.55 (1.29 - 1.87) | 190 | 37.34 (31.92 - 43.10) |
| Obese | 280 | 3.60 (3.08 - 4.20) | 310 | 37.75 (32.75 - 43.01) |
| **Waist circumference** |  |  |  |  |
| Very high | 420 | 3.08 (2.67 - 3.53) | 470 | 36.23 (31.97 - 40.72) |
| Other** | 160 | 1.08 (0.90 - 1.31) | 180 | 38.74 (33.19 - 44.59) |
| **Smoking status** |  |  |  |  |
| Current cigarette smoker | 110 | 2.63 (2.14 - 3.22) | 120 | 42.16 (35.54 - 49.07) |
| Ex-regular cigarette smoker | 190 | 1.88 (1.58 - 2.23) | 210 | 33.00 (28.07 - 38.32) |
| Never regular cigarette smoker | 310 | 1.85 (1.58 - 2.16) | 340 | 36.87 (32.32 - 41.67) |
| **Frequency of alcohol consumption** |  |  |  |  |
| At least once per week | 230 | 1.38 (1.18 - 1.62) | 260 | 40.39 (35.24 - 45.77) |
| Once or twice per month | 90 | 2.31 (1.85 - 2.89) | 100 | 41.80 (34.55 - 49.43) |
| Less than once or twice a month | 130 | 2.61 (2.17 - 3.14) | 150 | 37.10 (31.53 - 43.03) |
| Non-drinker/Not at all in last 12 months | 150 | 3.17 (2.55 - 3.93) | 170 | 31.06 (26.11 - 36.50) |

All estimates are weighted, and all are age-adjusted except where the predictor is age or sex or where there is no predictor (i.e., total population estimates).
\* A banded age category, 16-44 years, was required for undiagnosed diabetes due to small sample sizes in the youngest age groups. For non-diabetic hyperglycaemia we used 10-year age bands but also report estimates for the banded age group for comparability.
\*\* The Health Survey for England (HSE) uses thresholds to categorise waist circumference, with measurements above 102cm for males and 88cm for females considered very high.

After adjusting for age differences between groups, there was a higher prevalence of undiagnosed T2D among adults:

- From Black or Asian ethnic groups (5·02% (3·99-6·31) versus White-Mixed-Other 1·69% (1·48-1·93))
- Reporting no educational qualifications (2·60% (2·13-3·17) versus degree level or equivalent 1·48% (1·20-1·82))
- From ’routine and manual’ occupational classes (2·54% (2·15-2·99) versus ‘managerial and professional’ 1·43% (1·19-1·72))
- Reporting ‘bad or very bad’ health (3·11% (2·47-3·92) versus ‘very good or good’ 1·65% (1·44-1·88))
- Classed as ‘obese’ (BMI >30kg/m^2^), particularly among men (male: ‘obese’ 4·50% (3·68-5·50), ‘not overweight or obese’ 1·05% (0·71-1·56); female: ‘obese’ 1·27% (1·03-1·57), ‘not overweight or obese’ 0·34% (0·25-0·47)) (Table S4)
- With a ‘very high’ waist circumference (>102cm for males and >88cm for females), particularly among men (male: ‘very high’ 4·01% (3·31-4·85) versus ‘other’ 1·37% (1·08-1·74); female: ‘very high’ 1·10% (0·92-1·31) versus ‘other’ 0·34% (0·26-0·44)) (Table S4)
- Reporting no alcohol consumption in the past 12 months (3·17% (2·55-3·93) versus consumption at least once per week 1·38% (1·18-1·62)) (Table 2).

There were no differences in the age-adjusted prevalence of undiagnosed T2D by relationship status, region, rural-urban, antidepressant prescription, or smoking status.

### Percentage of adults with T2D who were undiagnosed

An estimated 30·21% (28·13-32·28) of adults with T2D were undiagnosed. Younger adults were more likely to be undiagnosed than older adults (50·03% (39·55-60·51) of those aged 16-44-years versus 27·43% (23·51-31·34) aged 75 and over) (Table 2).

After adjusting for age differences between groups, a higher proportion of adults with T2D were undiagnosed if they reported ‘very good or good’ general health (45·93% (40·97-50·98) versus ‘bad or very bad’ health (25·42% (20·38-31·23)).

Interaction effects were additionally found by sex for the following variables, with stratified results showing a higher proportion of women with T2D were undiagnosed if they:

- Were not prescribed antidepressants (22·58% (18·36-27·44)) versus prescribed antidepressants (12·91% (9·35-17·56))
- Were ‘not overweight or obese’ (51·24% (39·91-62·45) versus ‘obese’ (31·25% (25·64-37·47))
- Did not have a ‘very high’ waist circumference (30·99% (23·15-40·11)) versus ‘very high’ (17·75% (14·24-21·90)) (see Table S5).

There were no differences in the percentage of adults with T2D who were undiagnosed by sex, ethnicity, relationship status, region, rural-urban classification, education, socio-economic classification, smoking status, or alcohol consumption.

After adjusting for sociodemographic factors, younger adults, those living in the East Midlands, those with ‘very good or good’ general health, and those reporting more frequent alcohol consumption, were more likely to be undiagnosed (Figure 1). Interaction effects by sex were found for some additional predictors (Table S3) with stratified results showing that women who lived in rural areas, were not prescribed antidepressants, were ‘not overweight or obese’, or did not have a ‘very high’ waist circumference were more likely to be undiagnosed; these associations were not found for men (Table S5 and Figures S2-3). Women aged 55-64 were also more likely to be undiagnosed than those aged 75^+^.

**Figure 1.**
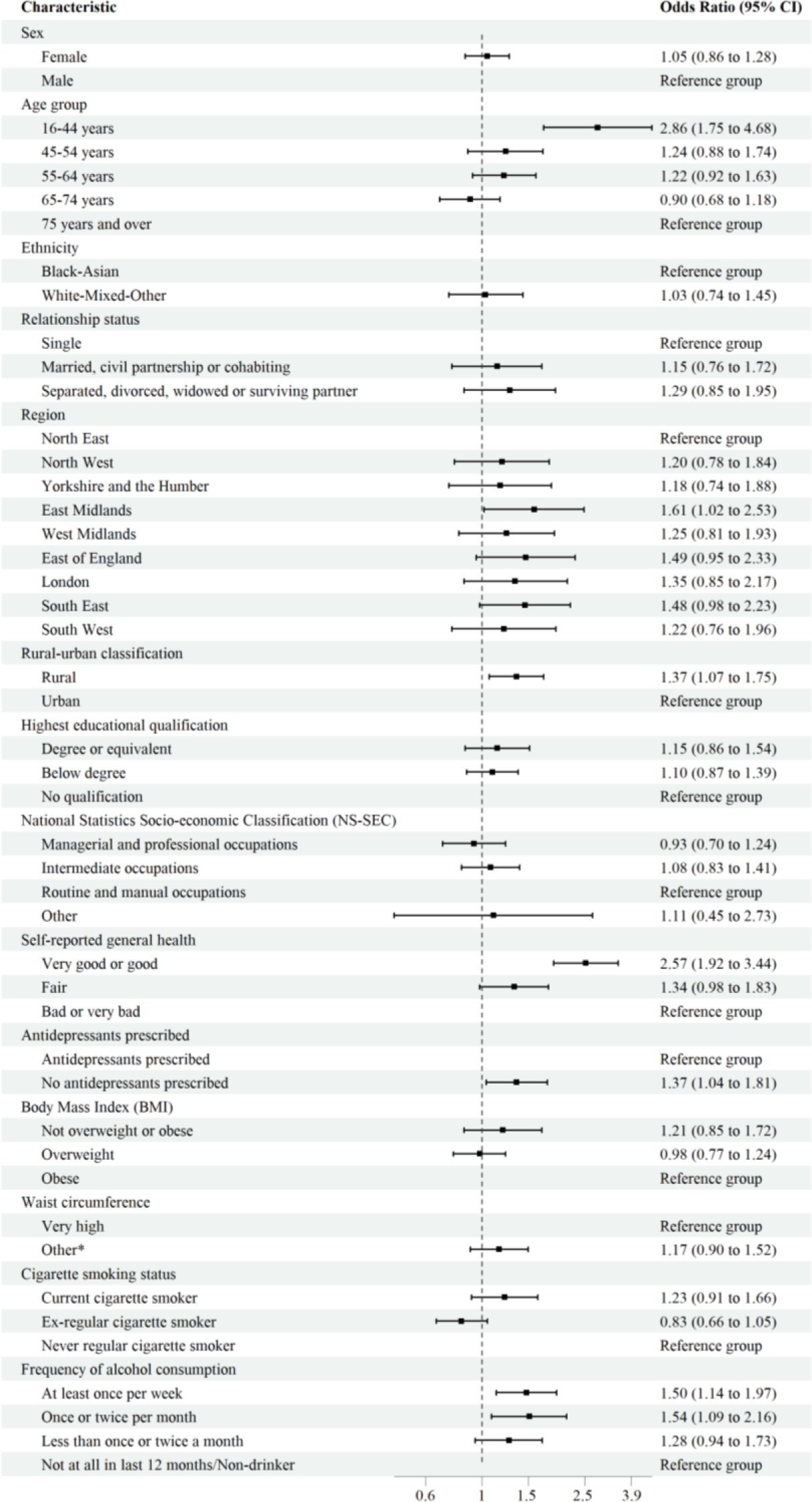
Adjusted odds ratios for undiagnosed type 2 diabetes, among those with type 2 diabetes, by characteristics Models were adjusted for age group, sex, ethnicity, relationship status, region, rural-urban classification, and highest educational qualification. * The Health Survey for England (HSE) uses thresholds to categorise waist circumference, with measurements above 102cm for males and 88cm for females considered very high.

### Non-diabetic hyperglycaemia (NDH)

An estimated 11·54% (11·11-11·97) of adults had NDH, equating to 5,121,800 (4,931,800-5,311,800) individuals living in private households in England. There was a higher prevalence of NDH among older adults (25·94% (24·26-27·62) of those aged 75 years and over compared with 3·96% (3·51-4·41) aged 16-44) (Table 3).

**Table 3.** Percentage with non-diabetic hyperglycaemia by characteristics.

|  | **Non-diabetic hyperglycaemia**<br>**(among all adults)** |  |
| --- | --- | --- |
|  | **Weighted**<br>**Number** | **Percentage**<br>**(95% CI)** |
| **Total** | 3090 | 11.54 (11.11 - 11.97) |
| **Sex** |  |  |
| Female | 1590 | 11.61 (11.06 - 12.16) |
| Male | 1500 | 11.46 (10.86 - 12.06) |
| **Age group** |  |  |
| 16-44 years* | 490 | 3.96 (3.51 - 4.41) |
| 16-24 years | 60 | 1.68 (0.94 - 2.42) |
| 25-34 years | 130 | 2.99 (2.30 - 3.69) |
| 35-44 years | 300 | 6.80 (5.96 - 7.65) |
| 45-54 years | 520 | 11.08 (10.13 - 12.03) |
| 55-64 years | 700 | 18.17 (16.99 - 19.35) |
| 65-74 years | 710 | 22.25 (21.00 - 23.50) |
| 75 years and over | 650 | 25.94 (24.26 - 27.62) |
| **Ethnicity** |  |  |
| Black-Asian | 400 | 21.71 (19.30 - 24.33) |
| White-Mixed-Other | 2690 | 9.92 (9.43 - 10.43) |
| **Relationship status** |  |  |
| Single | 300 | 10.56 (9.32 - 11.95) |
| Married, civil partnership or cohabiting | 2050 | 10.84 (10.24 - 11.46) |
| Separated, divorced, widowed, or surviving partner | 740 | 11.87 (10.92 - 12.89) |
| **Region** |  |  |
| North East | 160 | 10.91 (9.62 - 12.35) |
| North West | 370 | 10.13 (9.09 - 11.27) |
| Yorkshire and the Humber | 300 | 10.69 (9.48 - 12.04) |
| East Midlands | 310 | 12.64 (11.27 - 14.15) |
| West Midlands | 420 | 13.59 (12.18 - 15.14) |
| East of England | 330 | 10.34 (9.18 - 11.62) |
| London | 380 | 11.29 (9.86 - 12.89) |
| South East | 500 | 10.27 (9.37 - 11.24) |
| South West | 310 | 9.60 (8.53 - 10.79) |
| **Rural-urban classification** |  |  |
| Rural | 640 | 10.07 (9.27 - 10.94) |
| Urban | 2450 | 11.19 (10.61 - 11.79) |
| **Educational qualifications** |  |  |
| Degree or equivalent qualification | 580 | 8.66 (7.93 - 9.45) |
| Below degree qualification | 1520 | 11.23 (10.57 - 11.91) |
| No qualification | 970 | 13.48 (12.41 - 14.63) |
| **National Statistics Socio-economic Classification (NS-SEC)** |  |  |
| Managerial and professional occupations | 900 | 9.04 (8.40 - 9.72) |
| Intermediate occupations | 760 | 10.53 (9.75 - 11.37) |
| Routine and manual occupations | 1310 | 12.81 (12.01 - 13.66) |
| Other | 70 | 15.38 (11.02 - 21.06) |
| **Self-reported general health** |  |  |
| Very good or good | 2000 | 10.32 (9.79 - 10.88) |
| Fair | 760 | 13.12 (12.07 - 14.24) |
| Bad or very bad | 320 | 11.84 (10.41 - 13.43) |
| **Antidepressants prescribed** |  |  |
| Antidepressants prescribed | 430 | 12.50 (11.33 - 13.78) |
| No antidepressants prescribed | 2650 | 10.76 (10.22 - 11.33) |
| **Body Mass Index (BMI)** |  |  |
| Not overweight or obese | 660 | 8.28 (7.64 - 8.95) |
| Overweight | 1060 | 10.22 (9.53 - 10.96) |
| Obese | 1070 | 14.84 (13.77 - 15.98) |
| **Waist circumference** |  |  |
| Very high | 1760 | 13.54 (12.72 - 14.40) |
| Other** | 1240 | 8.91 (8.38 - 9.48) |
| **Smoking status** |  |  |
| Current cigarette smoker | 610 | 15.56 (14.33 - 16.88) |
| Ex-regular cigarette smoker | 940 | 10.10 (9.35 - 10.90) |
| Never regular cigarette smoker | 1540 | 10.08 (9.46 - 10.74) |
| **Frequency of alcohol consumption** |  |  |
| At least once per week | 1430 | 9.08 (8.55 - 9.64) |
| Once or twice per month | 380 | 10.95 (9.93 - 12.07) |
| Less than once or twice a month | 610 | 13.37 (12.21 - 14.63) |
| Non-drinker/Not at all in last 12 months | 670 | 15.42 (14.07 - 16.87) |

All estimates are weighted, and all are age-adjusted except where the predictor is age or sex or where there is no predictor (i.e., total population estimates).
\* A banded age category, 16-44 years, was required for undiagnosed diabetes due to small sample sizes in the youngest age groups. For non-diabetic hyperglycaemia we used 10-year age bands but also report estimates for the banded age group for comparability.
\*\* The Health Survey for England (HSE) uses thresholds to categorise waist circumference, with measurements above 102cm for males and 88cm for females considered very high.

After adjusting for age differences between groups, there was a higher prevalence of NDH among adults:

- From Black or Asian ethnic groups (21·71% (19·30-24·33) versus White, Mixed or Other (9·92% (9·43-10·43))
- Living in the East (13·59% (12·18-15·14)) or West Midlands (12·64% (11·27-14·15)) versus South West 9·60% (8·53-10·79))
- Reporting no qualifications (13·48% (12·41-14·63) versus degree level or equivalent (8·66% (7·93-9·45))
- From ‘routine and manual’ occupational classes (12·81% (12·01-13·66) versus ‘managerial and professional’ 9·04% (8·40-9·72))
- Reporting ‘fair’ (13·12% (12·07-14·24)) or ‘bad or very bad’ (11·84% (10·41-13·43)) health versus ‘very good or good’ (10·32% (9·79-10·88))
- Prescribed antidepressants (12·50% (11·33-13·78) versus 10·76% (10·22-11·33) not prescribed antidepressants)
- Classed as ‘obese’, particularly among women (female: ‘obese’ 14·61% (13·16-16·19), ‘not overweight or obese’ 7·77% (7·06-8·56); male: ‘obese’ 14·84% (13·77-15·98)), ‘not overweight or obese’ 8·28% (7·64-8·95)) (Table S6)
- With a ‘very high’ waist circumference, particularly among women (female: ‘very high’ 13·07% (12·01-14·21), ‘other’ 7·85% (7·21-8·54); male: (‘very high’ 13·54% (12·72-14·40), 8·91% (8·38-9·48) ‘other’)) (Table S6)
- Who currently smoke (15·56% (14·33-16·88)) versus ex-smokers (10·10% (9·35-10·90)) and those who never regularly smoked (10·08% (9·46-10·74))
- Reporting no alcohol consumption in the past 12 months (15·42% (14·07-16·87) versus consumption at least once per week (9·08% (8·55-9·64)) (Table 3).

There were no differences in prevalence of NDH by sex, relationship status or rural-urban.

After adjusting for sociodemographic factors, lower odds of NDH were found in younger adults, and higher odds among those from Black or Asian ethnic groups, those living in the East or West Midlands, those with fewer qualifications or belonging to routine and manual occupational classes, adults in ‘fair’ or ‘bad or very bad’ general health, those who are prescribed antidepressants, individuals with a higher BMI or higher waist circumference, current smokers, and those who report less frequent alcohol consumption (Figure 2). Women also had lower odds of NDH than men in adjusted models, and individuals who were separated/divorced/widowed had higher odds compared to those who were single.

**Figure 2.**
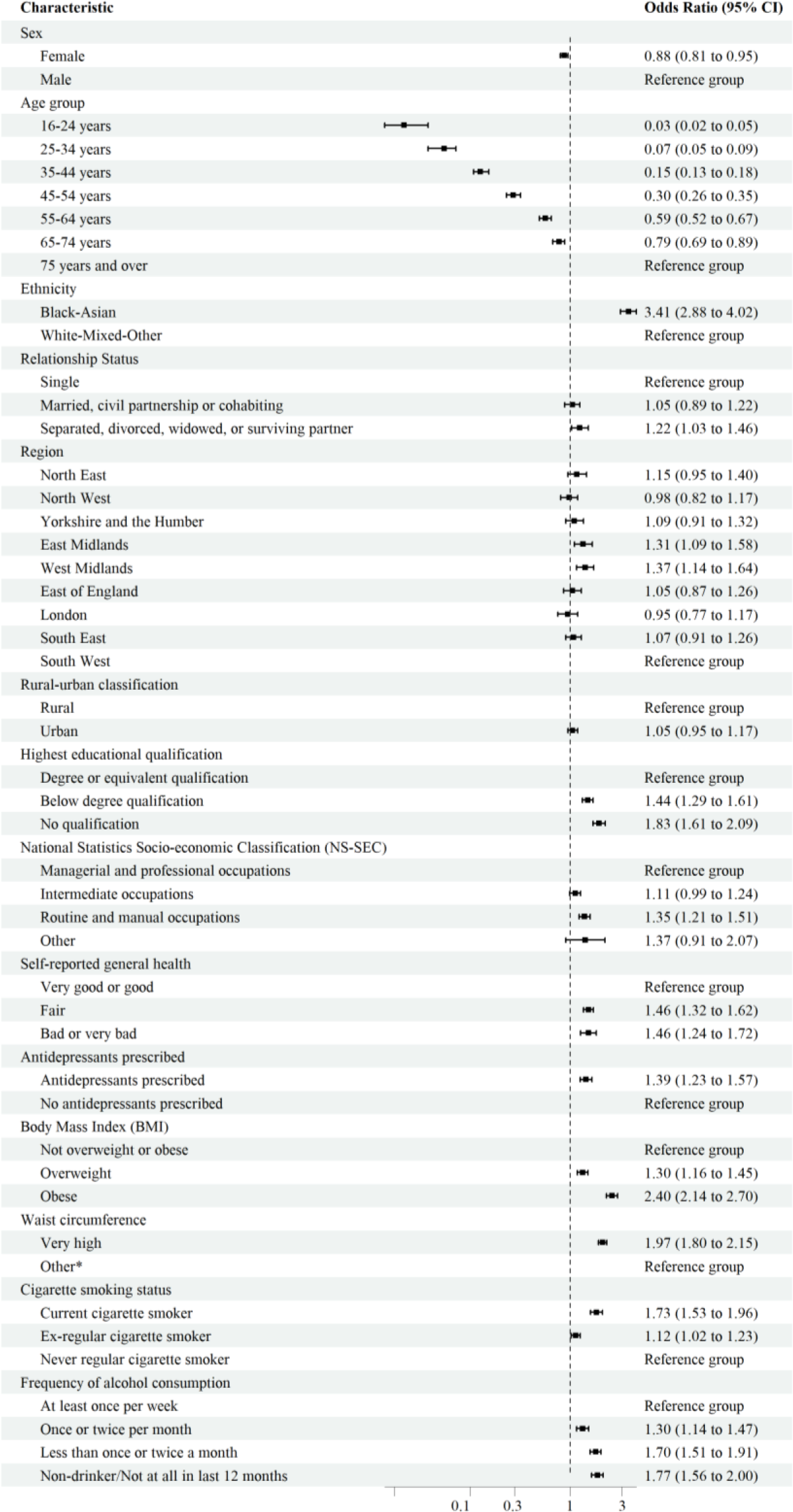
Adjusted odds ratios for non-diabetic hyperglycaemia, among adults with no evidence of diabetes, by characteristics. Models were adjusted for age group, sex, ethnicity, relationship status, region, rural-urban classification, and highest educational qualification. * The Health Survey for England (HSE) uses thresholds to categorise waist circumference, with measurements above 102cm for males and 88cm for females considered very high.

## Discussion

Between 2013-2019, an estimated 2·3% of adults in England had undiagnosed type 2 diabetes (T2D), representing 30·2% of those with T2D, and 11·5% had NDH. This equates to nearly 1 million adults with undiagnosed T2D and over 5 million with NDH. Risk factors for undiagnosed T2D aligned with known risk factors for T2D generally, with prevalence higher in males, older adults, those in manual and routine occupations, those with worse self-reported general health, and those with a higher BMI or waist circumference. Prevalence was more than double in those from Black and Asian ethnic groups compared with White, Mixed and Other ethnic groups.

Among those with T2D, younger adults and those self-reporting good health were proportionally more likely to be undiagnosed. Women, but not men, were more likely to be undiagnosed if they were not prescribed antidepressants, were not overweight or obese, and did not have a ‘very high’ waist circumference. These individuals may be less likely to be identified by primary care. Screening and awareness campaigns likely focus on populations known to have higher risk of T2D, thereby increasing the likelihood of diagnosis. Groups of known increased risk or with family history may also have greater awareness of T2D, increasing likelihood of diagnosis. Our previous work similarly showed that among those with hypertension, individuals with lower risk profile were more likely to be undiagnosed.^19^

The observed differences in the likelihood of T2D being undiagnosed by BMI and waist circumference for women but not men suggest that current systems for detection and diagnosis are performing poorly among women if they have a lower BMI. Sex-based differences in T2D onset exist, with prevalence higher in men during midlife, and women experiencing onset at a higher age-adjusted average BMI.^17^ Known sex-based differences may impact clinical risk assessment, reducing likelihood of testing in women of healthier BMI. Women living in rural areas also had higher odds of being undiagnosed compared with women in urban areas, likely related to healthcare access, although it is unclear why this would not equally apply to men.

In adjusted models, individuals with T2D living in the East Midlands had the highest odds of being undiagnosed, while those living in the East/West Midlands had highest odds of NDH. Between April 2013 and July 2022 NHS care was controlled by clinical commissioning groups,^20^ with a variety of regional factors impacting care and potentially contributing to disparities in diagnosis. This may be exacerbated by regional variation in healthcare engagement, supported by regional differences in uptake of the NHS Health Check.^21^ However, there may be residual confounding in our models, particularly for predictors like ethnicity where sample sizes necessitated a high degree of aggregation.

Older adults, Black and Asian ethnic groups, manual and routine occupational groups, those with higher BMI and waist circumference, and smokers also had higher NDH prevalence which aligns with previous literature.^13,14^ However, the sizeable NDH prevalence across all groups is noteworthy, including those typically considered “low risk”, such as younger adults (4·0% of 16–44-year-olds) and those who were not overweight or obese (8·3%). As NDH prevalence increases in England,^14,22^ we provide additional evidence to support identification of the expanding groups at high risk of developing T2D. Our results highlight a need to consider primary prevention across the population, including in those with lower T2D risk and in those without a diabetes diagnosis.

The HSE provides a unique opportunity to explore sociodemographic and health-related risk factors for NDH and undiagnosed T2D. Stringent protocols for accurate and standardised HbA1c measurements and robust survey design and weighting, allowed us to produce estimates representative of adults in England. We explored a wide variety of health and sociodemographic predictors for the first time, providing novel insights into groups most at risk, and we provided some of the first insights into differences in risk factors by sex. Unlike many previous studies, we excluded individuals with type 1 diabetes since the risk profiles and population prevalence of type 1 and T2D differ. There was minimal missing data within our analysis.

However, there remain several limitations. Despite boosting sample size by pooling survey years, we observed small numbers for undiagnosed T2D and some predictor groups. This mainly affected younger adults and Black and Asian ethnic groups, particularly in sex-stratified results. Our analysis relied on self-reports of diagnosis, which are unlikely to completely align with clinical diagnoses. We used age at diagnosis to classify the 93 individuals who did not know their diabetes type, but we cannot be certain this correctly excluded all individuals with type 1 diabetes and included all with T2D. The impact of this would be minimal considering the small number of individuals affected (0·3% of the sample). Elevated HbA1c at a single point in time is not equivalent to a medical diagnosis; NICE guidance recommends confirming with a repeat test and further monitoring.^23^ COVID-19 has impacted prevalence and diagnosis of T2D;^12^ future research should make use of post-pandemic data once available to assess potential change in risk factors.

In England, the NHS Health Check is offered every 5 years to adults aged 40-74 years without certain pre-existing conditions, culminating in a risk assessment score for conditions including diabetes and NDH, which shape discussions on modifiable risk factors. Uptake of the Health Check was estimated at 52·6% of those eligible between 2012-2017, with regional variation.^21^ There was a reduction to 38·9% uptake in 2022/23, however, this occurred alongside an increase in the total number of Health Checks offered.^24^ Diabetes screening is offered by some pharmacies, providing a risk assessment score and recommendation for further testing in those deemed high risk. However, NDH and T2D are frequently asymptomatic and are often diagnosed indirectly though concerns about another condition. Campaigns such as “Healthier You” by the NDH Diabetes Prevention Programme (NDPP), and “Know Your Risk” by Diabetes UK aim to increase awareness and help identify those at risk of developing T2D. Recent analysis of the NDPP showed reduced incidence of T2D.^25^ Between 2016-2017 uptake was around 56% with 34% achieving the required completion, and variation by sociodemographic factors.^26^ Evidence shows improved completion when including the COVID-19 pandemic, when remote participation was made available.^27^ Our data suggest that around 1 in 9 adults in England have NDH and future strategies should include improving provision across all population groups.

This study builds on existing evidence on risk factors for NDH and undiagnosed T2D in England. Risk factors aligned with known risks for T2D generally, but some groups less likely to have T2D were more likely to be undiagnosed if they did have the condition, and considerable prevalence was found in groups typically considered “low risk”. Findings highlight the burden of T2D and NDH in England and emphasise the need for awareness of risk across all groups. Policy and public health campaigns should consider these findings in their design.

## Data Availability

Data from the Health Survey for England is available for researchers to access via the UK Data Service.

## Contribution of authors

EC, KF, and EM conceived the idea for the study. Study design was led by EC, KF, and EM. EC and EM were responsible for curating and cleaning the data. Analysis was carried out by EC, EM, and CK-H. EC and KF had leadership responsibility and supervised the project. Methodological advice was provided by VN, NB, and MW. Topic and policy expertise were provided by KK, MW, NB, RJ, and CS. EC, EM, CK-H, and KF wrote the first draft of the publication. All authors reviewed and commented on the publication and approved the final version. EC, EM, and KF accessed and verified the underlying data.

## Acknowledgements

KK is supported by the National Institute for Health Research (NIHR) Applied Research Collaboration East Midlands (ARC EM) and the NIHR Leicester Biomedical Research Centre (BRC).

## Data sharing agreement

Data from the Health Survey for England is available for researchers to access via the UK Data Service. Methods reports and related documents for the Health Survey are available on the NHD Digital website.

