## Supplementary material for "Sociodemographic and health-related differences in non-diabetic hyperglycaemia and undiagnosed type 2 diabetes in the Health Survey for England 2013-2019"

Figure S1. Flow chart of sample selection

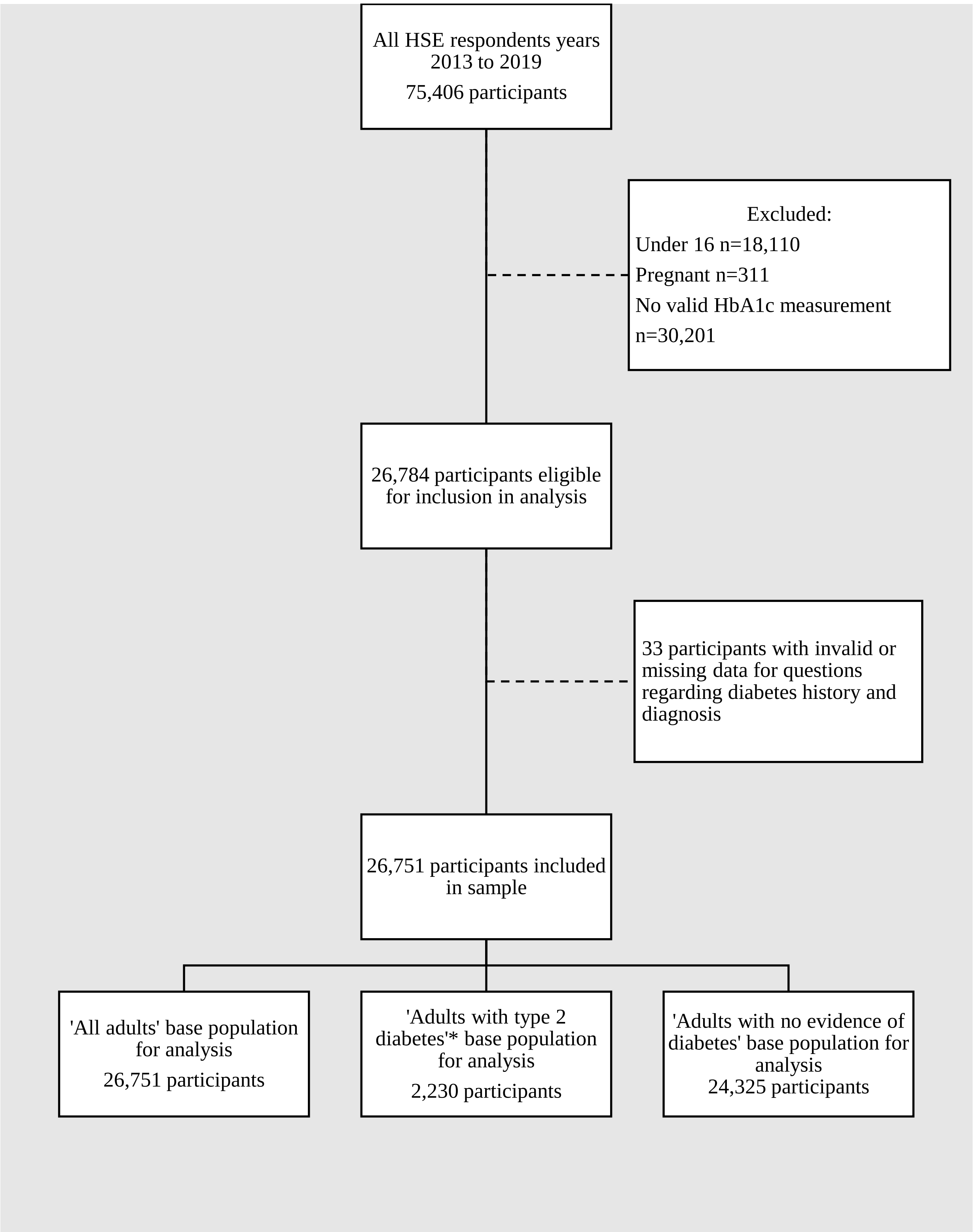


* ‘Adults with type 2 diabetes’ base population includes only those with type 2 diabetes· 196 individuals in the sample had type 1 or type ‘unknown’ diabetes and are only present in the ‘all adults’ base population·

Table S1 Predictors included in analysis

| Characteristic | Levels | Details |
|---|---|---|
| **Sex** | Male<br>Female | Self-reported binary variable |
| **Age group** | 16-24 years<br>25-34 years<br>35-44 years<br>45-54 years<br>55-64 years<br>65-74 years<br>75 years and over | Derived from self-reported age at last birthday; aggregated age group (16-44 years) used where sample size necessitated |
| **Ethnicity** | Black-Asian<br>White-Mixed-Other | Derived from respondent ethnicity selection from 17 ethnic group options |
| **Relationship status** | Single<br>Married, civil partnership or cohabiting<br>Separated, divorced, widowed, or surviving partner | Derived from responses to interview questions about relationships within the household |
| **Region** | North East<br>North West<br>Yorkshire and the Humber<br>East Midlands<br>West Midlands<br>East of England<br>London<br>South East<br>South West | Defined by household address |
| **Rural-urban classification** | Rural<br>Urban | Derived variable on rurality of dwelling· Rural includes town/fringe/village, hamlet and isolated dwellings |
| **Highest educational qualification** | Degree or equivalent qualification<br>Below degree qualification<br>No qualification | Derived from self-reports of completed qualifications from a list of 29 options |
| **National Statistics Socio-economic Classification (NS-SEC)** | Managerial and professional occupations<br>Intermediate occupations<br>Routine and manual occupations<br>Other<br>Not applicable | Classification derived from responses to questions about employment status and working patterns |
| **Self-reported general health** | Very good or good<br>Fair<br>Bad or very bad | Derived from respondent self-assessment of general health on a scale from 1 (very good) to 5 (very bad) |
| **Antidepressants prescribed** | Antidepressants prescribed<br>No antidepressants prescribed | Derived from participant provided list of medications currently prescribed |
| **Body Mass Index (BMI)** | Not overweight or obese | |

| | | |
|---|---|---|
| | Overweight<br>Obese | Calculated from height and weight measurements taken by the interviewer, or estimated weight if participants were believed to be above 130kg; BMI <25 not overweight or obese; 25 to <30 overweight; ≥30 obese |
| **Waist circumference** | Very high waist circumference<br>(>102cm males/>88cm females)<br>Other waist measurement<br>Not applicable | Derived from waist measurements taken by the interviewer and categorisation based on waist circumference thresholds from NHS Digital guidelines |
| **Smoking status** | Current cigarette smoker<br>Ex-regular cigarette smoker<br>Never regular cigarette smoker | Derived from responses to several questions about current and past smoking habits |
| **Frequency of alcohol consumption** | At least once per week<br>Once or twice per month<br>Infrequently<br>Non-drinker/Not at all in last 12 months | Derived from responses to several questions regarding alcohol consumption in past 12 months |

Table S2. Proportion of missing data

| | Missing data N (%) |
|---|---|
| **Sex** | 0 (0·00%) |
| **Age group** | 0 (0·00%) |
| **Ethnicity** | 17 (0·06%) |
| **Relationship status** | <10 (0·01%) |
| **Region** | 0 (0·00%) |
| **Rural-urban classification** | <10 (0·01%) |
| **Highest educational qualification** | 34 (0·13%) |
| **NS-SEC** | 12 (0·04%) |
| **Self-reported general health** | <10 (0·02%) |
| **Antidepressants prescribed** | 0 (0%) |
| **BMI** | 2,310 (8·64%) |
| **Waist circumference** | 0 (0%) |
| **Smoking status** | 46 (0·17%) |
| **Frequency of alcohol consumption** | 50 (0·19%) |

BMI – Body Mass Index; NS-SEC - National Statistics Socio-economic Classification (NS-SEC)

Table S3. Results of Wald test for sex interaction for each outcome and characteristic

| Outcome group | Characteristic | *F*-Statistic | *P*-value |
|---|---|---|---|
| Undiagnosed type 2 diabetes (All adults base population) | Age group | 3·10 | 0·0050 |
| | Ethnicity | 0·06 | 0·80 |
| | Relationship status | 1·80 | 0·16 |
| | Region | 0·69 | 0·70 |
| | Rural-urban classification | 3·25 | 0·072 |
| | Highest educational qualification | 2·39 | 0·092 |
| | NS-SEC | 1·78 | 0·13 |
| | Self-reported general health | 1·31 | 0·27 |
| | Antidepressants prescribed | 1·07 | 0·30 |
| | BMI | 7·57 | 0·00052 |
| | Waist circumference | 13·26 | <0·0001 |
| | Smoking status | 0·02 | 0·98 |
| | Frequency of alcohol consumption | 2·48 | 0·059 |
| Undiagnosed type 2 diabetes (Adults with type 2 diabetes base population) | Age group | 17·98 | <0·0001 |
| | Ethnicity | 1·56 | 0·21 |
| | Relationship status | 1·75 | 0·17 |
| | Region | 0·66 | 0·72 |
| | Rural-urban classification | 4·12 | 0·043 |
| | Highest educational qualification | 1·48 | 0·23 |
| | NS-SEC | 0·82 | 0·51 |
| | Self-reported general health | 0·71 | 0·49 |
| | Antidepressants prescribed | 6·47 | 0·011 |
| | BMI | 5·16 | 0·0059 |
| | Waist circumference | 4·64 | 0·0098 |
| | Smoking status | 2·72 | 0·066 |
| | Frequency of alcohol consumption | 0·90 | 0·44 |
| Non-diabetic hyperglycaemia (All adults base population) | Age group | 2·65 | 0·014 |
| | Ethnicity | 0·00042 | 0·98 |
| | Relationship status | 1·84 | 0·16 |
| | Region | 1·10 | 0·36 |
| | Rural-urban classification | 4·66 | 0·031 |
| | Highest educational qualification | 1·94 | 0·14 |
| | NS-SEC | 1·92 | 0·10 |
| | Self-reported general health | 1·15 | 0·32 |
| | Antidepressants prescribed | 0·28 | 0·59 |
| | BMI | 7·32 | 0·00067 |
| | Waist circumference | 12·02 | <0·0001 |
| | Smoking status | 0·03 | 0·97 |
| | Frequency of alcohol consumption | 1·68 | 0·17 |

BMI – Body Mass Index; NS-SEC - National Statistics Socio-economic Classification (NS-SEC)

Table S4. Percentage of all adults with undiagnosed type 2 diabetes by sex and characteristics

| | Undiagnosed diabetes (among all females) | | Undiagnosed diabetes (among all males) | |
|---|---|---|---|---|
| | Weighted Number | Percentage (95% CI) | Weighted Number | Percentage (95% CI) |
| **Total** | 270 | 1·94 (1·72 - 2·17) | 340 | 2·57 (2·25 - 2·90) |
| **Age group** | | | | |
| 16-44 years | 30 | 0·41 (0·24 – 0·58) | 60 | 0·99 (0·58 – 1·39) |
| 45-54 years | 40 | 1·87 (1·35 - 2·39) | 70 | 3·13 (2·32 - 3·93) |
| 55-64 years | 60 | 3·11 (2·39 - 3·82) | 80 | 3·99 (2·98 - 4·99) |
| 65-74 years | 60 | 3·48 (2·71 - 4·24) | 60 | 4·12 (3·26 - 4·97) |
| 75 years and over | 80 | 5·39 (4·23 - 6·54) | 60 | 5·75 (4·41 - 7·08) |
| **Ethnicity** | | | | |
| Black-Asian | 40 | 1·87 (1·34 - 2·61) | 60 | 6·05 (4·44 - 8·19) |
| White-Mixed-Other | 230 | 0·63 (0·53 - 0·74) | 270 | 2·02 (1·70 - 2·39) |
| **Relationship status** | | | | |
| Single | 20 | 0·73 (0·49 - 1·09) | 50 | 2·41 (1·70 - 3·41) |
| Married, civil partnership or cohabiting | 140 | 0·65 (0·53 - 0·79) | 240 | 2·35 (1·97 - 2·81) |
| Separated, divorced, widowed, or surviving partner | 110 | 1·00 (0·79 - 1·27) | 50 | 2·45 (1·83 - 3·26) |
| **Region** | | | | |
| North East | 10 | 0·66 (0·43 - 1·02) | 10 | 2·03 (1·36 - 3·02) |
| North West | 30 | 0·65 (0·45 - 0·92) | 30 | 1·81 (1·23 - 2·65) |
| Yorkshire and the Humber | 30 | 0·82 (0·58 - 1·16) | 30 | 2·25 (1·52 - 3·31) |
| East Midlands | 30 | 1·00 (0·70 - 1·42) | 30 | 2·37 (1·65 - 3·39) |
| West Midlands | 30 | 0·75 (0·51 - 1·10) | 50 | 2·88 (2·10 - 3·95) |
| East of England | 20 | 0·58 (0·40 - 0·84) | 40 | 2·41 (1·71 - 3·39) |
| London | 30 | 0·78 (0·54 - 1·12) | 50 | 3·00 (2·05 - 4·37) |
| South East | 40 | 0·70 (0·53 - 0·92) | 60 | 2·58 (1·80 - 3·69) |
| South West | 30 | 0·67 (0·44 - 1·03) | 30 | 1·67 (0·98 - 2·82) |
| **Rural-urban classification** | | | | |
| Rural | 70 | 0·84 (0·65 - 1·09) | 60 | 1·90 (1·37 - 2·63) |
| Urban | 200 | 0·70 (0·59 - 0·83) | 280 | 2·49 (2·12 - 2·93) |
| **Educational qualifications** | | | | |
| Degree or equivalent qualification | 30 | 0·45 (0·31 - 0·63) | 70 | 1·94 (1·50 - 2·49) |
| Below degree qualification | 120 | 0·73 (0·60 - 0·87) | 170 | 2·46 (2·02 - 2·99) |
| No qualification | 110 | 1·12 (0·87 - 1·43) | 90 | 2·74 (2·03 - 3·70) |
| **National Statistics Socio-economic Classification (NS-SEC)** | | | | |
| Managerial and professional occupations | 50 | 0·50 (0·38 – 0·65) | 100 | 1·74 (1·39 – 2·17) |
| Intermediate occupations | 70 | 0·65 (0·50 – 0·84) | 70 | 2·29 (1·69 – 3·10) |

| | | | | |
|---|---|---|---|---|
| Routine and manual occupations | 120 | 0·89 (0·73 – 1·09) | 160 | 3·10 (2·50 – 3·84) |
| Other | 10 | 1·93 (1·07 – 3·44) | - | - |
| **Self-reported general health** | | | | |
| Very good or good | 140 | 0·61 (0·51 - 0·72) | 200 | 1·96 (1·65 - 2·33) |
| Fair | 80 | 0·98 (0·74 - 1·28) | 100 | 3·57 (2·68 - 4·75) |
| Bad or very bad | 50 | 1·23 (0·89 - 1·69) | 40 | 3·63 (2·60 - 5·06) |
| **Antidepressants prescribed** | | | | |
| Antidepressants prescribed | 50 | 0·77 (0·57 - 1·03) | 50 | 4·28 (3·12 - 5·85) |
| No antidepressants prescribed | 220 | 0·72 (0·61 - 0·85) | 290 | 2·22 (1·88 - 2·61) |
| **Body Mass Index (BMI)** | | | | |
| Not overweight or obese | 40 | 0·34 (0·25 - 0·47) | 40 | 1·05 (0·71 - 1·56) |
| Overweight | 70 | 0·59 (0·45 - 0·77) | 100 | 1·76 (1·38 - 2·25) |
| Obese | 120 | 1·27 (1·03 - 1·57) | 160 | 4·50 (3·68 - 5·50) |
| **Waist circumference** | | | | |
| Very high | 200 | 1·10 (0·92 - 1·31) | 220 | 4·01 (3·31 - 4·85) |
| Other* | 50 | 0·34 (0·26 - 0·44) | 110 | 1·37 (1·08 - 1·74) |
| **Smoking status** | | | | |
| Current cigarette smoker | 40 | 0·85 (0·63 - 1·14) | 70 | 3·40 (2·61 - 4·41) |
| Ex-regular cigarette smoker | 80 | 0·76 (0·60 - 0·96) | 110 | 2·09 (1·67 - 2·62) |
| Never regular cigarette smoker | 150 | 0·71 (0·59 - 0·87) | 150 | 2·20 (1·78 - 2·73) |
| **Frequency of alcohol consumption** | | | | |
| At least once per week | 70 | 0·40 (0·32 - 0·51) | 160 | 1·83 (1·51 - 2·22) |
| Once or twice per month | 40 | 0·81 (0·59 - 1·11) | 50 | 3·01 (2·20 - 4·10) |
| Infrequently | 80 | 1·15 (0·91 - 1·45) | 50 | 2·71 (1·99 - 3·68) |
| Non-drinker/Not at all in last 12 months | 80 | 1·17 (0·90 - 1·53) | 70 | 4·15 (3·01 - 5·70) |

All estimates are weighted, and all are age-adjusted except where the predictor is age or sex or where there is no predictor (i.e., total population estimates). Suppression has been used where estimates were based on a small number of respondents (<10) due to low reliability of estimates and to maintain confidentiality.

* The Health Survey for England (HSE) uses thresholds to categorise waist circumference, with measurements above 102cm for males and 88cm for females considered very high.

Table S5. Percentage of adults with type 2 diabetes who were undiagnosed, by sex and characteristics

| | Undiagnosed diabetes (among females with diabetes) | | Undiagnosed diabetes (among males with diabetes) | |
|---|---|---|---|---|
| | **Weighted Number** | **Percentage (95% CI)** | **Weighted Number** | **Percentage (95% CI)** |
| **Total** | 300 | 30·23 (27·30 – 33·15) | 380 | 30·19 (27·29 – 33·10) |
| **Age group** | | | | |
| 16-44 years | 30 | 41·50 (27·74 – 55·25) | 70 | 54·62 (40·77 – 68·48) |
| 45-54 years | 50 | 28·69 (21·34 - 36·03) | 80 | 32·76 (25·40 - 40·12) |
| 55-64 years | 70 | 34·35 (27·94 - 40·75) | 80 | 28·25 (22·48 - 34·02) |
| 65-74 years | 60 | 28·33 (22·94 - 33·72) | 70 | 22·54 (18·37 - 26·71) |
| 75 years and over | 90 | 27·39 (22·08 - 32·70) | 70 | 27·47 (21·83 - 33·11) |
| **Ethnicity** | | | | |
| Black-Asian | 40 | 17·76 (12·15 - 25·22) | 70 | 38·66 (29·06 - 49·23) |
| White-Mixed-Other | 260 | 20·65 (16·79 - 25·13) | 310 | 38·21 (32·54 - 44·22) |
| **Relationship status** | | | | |
| Single | 20 | 16·11 (10·08 - 24·75) | 50 | 35·41 (26·00 - 46·11) |
| Married, civil partnership or cohabiting | 160 | 19·79 (15·72 - 24·61) | 270 | 39·29 (33·02 - 45·94) |
| Separated, divorced, widowed, or surviving partner | 120 | 22·63 (17·36 - 28·94) | 60 | 37·65 (28·65 - 47·61) |
| **Region** | | | | |
| North East | 10 | 15·62 (9·81 - 23·97) | 20 | 33·23 (22·47 - 46·06) |
| North West | 30 | 18·51 (12·98 - 25·70) | 40 | 35·26 (25·79 - 46·05) |
| Yorkshire and the Humber | 30 | 20·52 (13·97 - 29·10) | 30 | 35·59 (24·61 - 48·32) |
| East Midlands | 40 | 26·08 (18·20 - 35·86) | 30 | 38·68 (27·91 - 50·68) |
| West Midlands | 30 | 17·22 (11·73 - 24·57) | 50 | 38·48 (29·05 - 48·87) |
| East of England | 30 | 19·56 (12·87 - 28·59) | 40 | 46·21 (35·52 - 57·27) |
| London | 40 | 16·87 (10·93 - 25·14) | 60 | 39·03 (29·53 - 49·45) |
| South East | 50 | 23·36 (17·17 - 30·95) | 70 | 39·50 (30·42 - 49·36) |
| South West | 30 | 21·83 (14·57 - 31·38) | 30 | 35·47 (24·76 - 47·86) |
| **Rural-urban classification** | | | | |
| Rural | 80 | 28·46 (21·77 - 36·26) | 70 | 40·86 (32·75 - 49·51) |
| Urban | 220 | 18·49 (14·97 - 22·62) | 310 | 37·72 (31·80 - 44·03) |
| **Educational qualifications** | | | | |
| Degree or equivalent qualification | 40 | 20·13 (14·13 - 27·84) | 80 | 40·34 (31·16 - 50·25) |
| Below degree qualification | 130 | 20·01 (15·87 - 24·92) | 190 | 39·67 (33·54 - 46·14) |
| No qualification | 130 | 19·69 (14·88 - 25·59) | 100 | 33·44 (26·46 - 41·23) |
| **National Statistics Socio-economic Classification (NS-SEC)** | | | | |
| Managerial and professional occupations | 60 | 22·57 (16·94 - 29·41) | 110 | 36·28 (28·79 - 44·52) |
| Intermediate occupations | 80 | 22·70 (17·08 - 29·50) | 80 | 38·36 (30·52 - 46·87) |

| | | | | |
|---|---|---|---|---|
| Routine and manual occupations | 140 | 18·82 (14·77 - 23·67) | 180 | 39·31 (32·65 - 46·38) |
| Other | 10 | 19·35 (10·03 - 34·05) | - | - |
| **Self-reported general health** | | | | |
| Very good or good | 160 | 28·98 (23·43 - 35·23) | 220 | 45·25 (38·18 - 52·53) |
| Fair | 90 | 15·73 (11·90 - 20·52) | 110 | 32·81 (26·47 - 39·85) |
| Bad or very bad | 50 | 12·30 (8·61 - 17·28) | 50 | 27·50 (19·91 - 36·65) |
| **Antidepressants prescribed** | | | | |
| Antidepressants prescribed | 50 | 12·91 (9·35 - 17·56) | 60 | 40·12 (30·20 - 50·91) |
| No antidepressants prescribed | 250 | 22·58 (18·36 - 27·44) | 320 | 38·01 (32·41 - 43·94) |
| **Body Mass Index (BMI)** | | | | |
| Not overweight or obese | 40 | 51·24 (39·91 - 62·45) | 40 | 33·96 (23·77 - 45·9) |
| Overweight | 80 | 33·26 (26·30 - 41·04) | 110 | 39·47 (32·37 - 47·04) |
| Obese | 140 | 31·25 (25·64 - 37·47) | 180 | 41·79 (34·70 - 49·23) |
| **Waist circumference** | | | | |
| Very high | 220 | 17·75 (14·24 - 21·90) | 240 | 40·06 (33·70 - 46·78) |
| Other* | 50 | 30·99 (23·15 - 40·11) | 120 | 35·92 (29·30 - 43·12) |
| **Smoking status** | | | | |
| Current cigarette smoker | 40 | 20·04 (14·32 - 27·31) | 80 | 48·14 (38·81 - 57·60) |
| Ex-regular cigarette smoker | 90 | 19·91 (15·16 - 25·70) | 120 | 32·98 (26·10 - 40·67) |
| Never regular cigarette smoker | 170 | 20·46 (16·28 - 25·40) | 170 | 37·45 (31·20 - 44·15) |
| **Frequency of alcohol consumption** | | | | |
| At least once per week | 80 | 22·43 (17·10 - 28·84) | 180 | 42·92 (36·08 - 50·03) |
| Once or twice per month | 40 | 24·58 (17·53 - 33·32) | 60 | 42·66 (32·29 - 53·71) |
| Infrequently | 90 | 22·94 (17·73 - 29·14) | 60 | 33·10 (24·76 - 42·65) |
| Non-drinker/Not at all in last 12 months | 90 | 16·06 (12·12 - 20·97) | 80 | 32·27 (24·83 - 40·74) |

All estimates are weighted, and all are age-adjusted except where the predictor is age or sex or where there is no predictor (i.e., total population estimates). Suppression has been used where estimates were based on a small number of respondents (<10) due to low reliability of estimates and to maintain confidentiality.

* The Health Survey for England (HSE) uses thresholds to categorise waist circumference, with measurements above 102cm for males and 88cm for females considered very high.

Figure S2. Adjusted odds ratios for undiagnosed type 2 diabetes, among females with type 2 diabetes, by characteristics

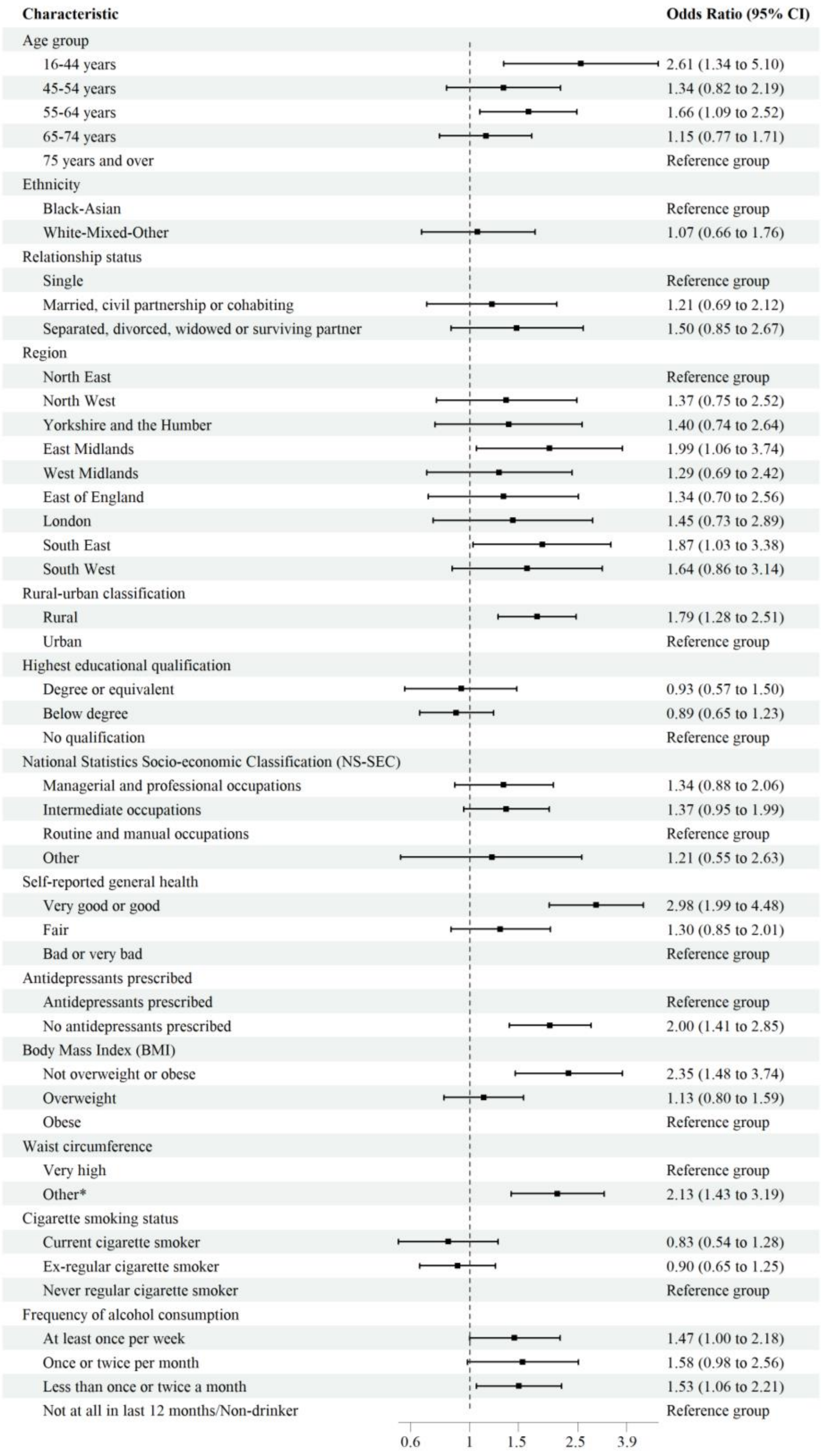


Models were adjusted for age, ethnicity, relationship status, region, rural-urban classification, and highest educational qualification.

* The Health Survey for England (HSE) uses thresholds to categorise waist circumference, with measurements above 102cm for males and 88cm for females considered very high.

Figure S3. Adjusted odds ratios for undiagnosed type 2 diabetes, among males with type 2 diabetes, by characteristics

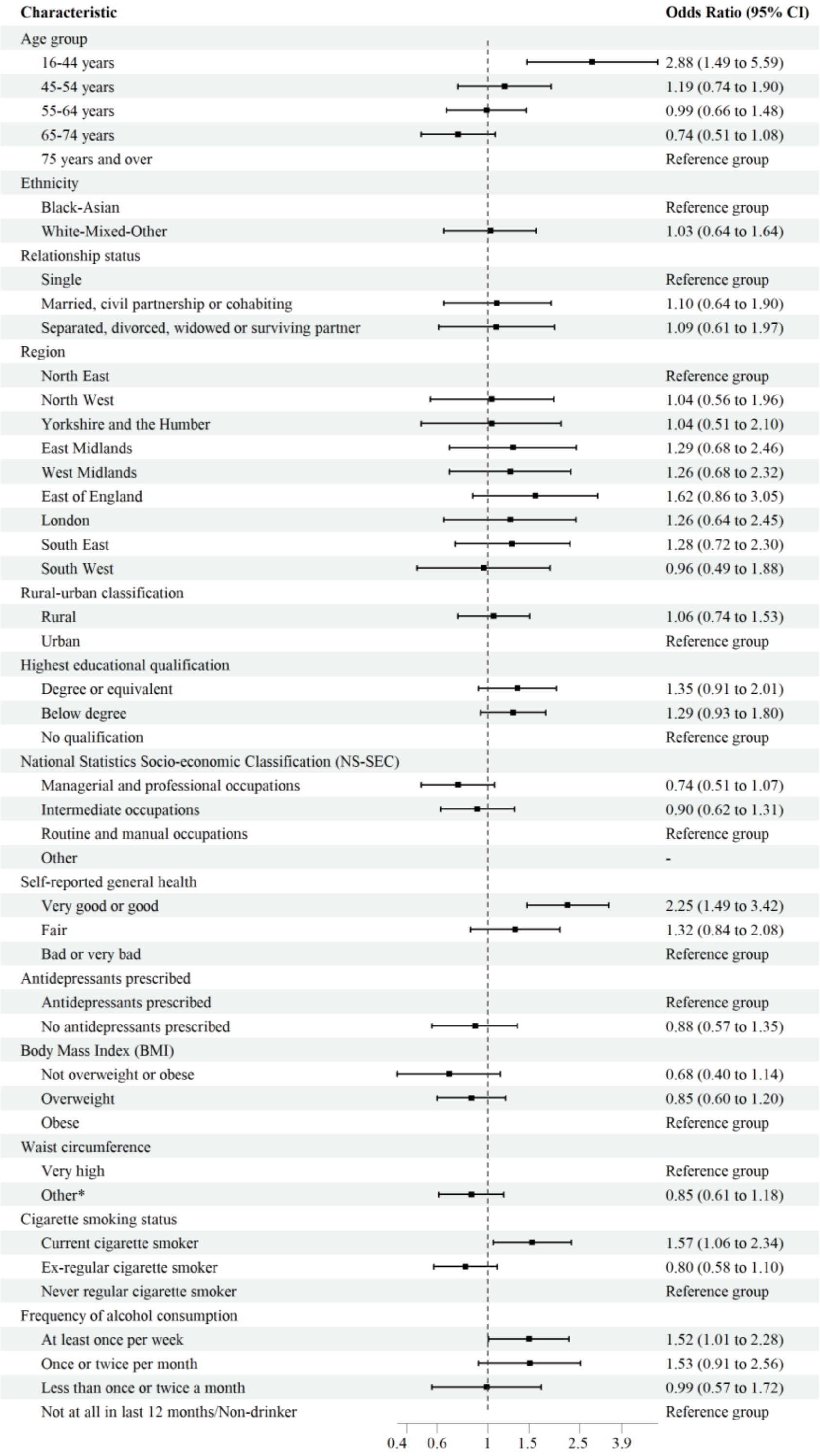


Models were adjusted for age, ethnicity, relationship status, region, rural-urban classification, and highest educational qualification.

* The Health Survey for England (HSE) uses thresholds to categorise waist circumference, with measurements above 102cm for males and 88cm for females considered very high.

Table S6. Percentage of all adults with non-diabetic hyperglycaemia (NDH) by sex and characteristics

| | Non-diabetic hyperglycaemia (among all females) | | Non-diabetic hyperglycaemia (among all males) | |
|---|---|---|---|---|
| | Weighted Number | Percentage (95% CI) | Weighted Number | Percentage (95% CI) |
| **Total** | 1590 | 11·61 (11·06 - 12·16) | 1500 | 11·46 (10·86 - 12·06) |
| **Age group** | | | | |
| 16-44 years* | 200 | 3·29 (2·74 – 3·84) | 290 | 4·63 (3·95 – 5·31) |
| 16-24 years | 30 | 1·59 (0·61 - 2·57) | 30 | 1·77 (0·66 - 2·89) |
| 25-34 years | 60 | 2·67 (1·87 - 3·48) | 70 | 3·32 (2·28 - 4·36) |
| 35-44 years | 120 | 5·29 (4·30 - 6·27) | 180 | 8·33 (7·00 - 9·67) |
| 45-54 years | 230 | 9·82 (8·64 – 11·00) | 290 | 12·37 (10·87 - 13·86) |
| 55-64 years | 380 | 19·03 (17·42 - 20·64) | 330 | 17·27 (15·62 - 18·93) |
| 65-74 years | 390 | 23·71 (22·00 - 25·43) | 320 | 20·69 (18·91 - 22·46) |
| 75 years and over | 380 | 26·55 (24·28 - 28·83) | 280 | 25·14 (22·77 - 27·52) |
| **Ethnicity** | | | | |
| Black-Asian | 200 | 22·20 (19·10 - 25·65) | 200 | 21·22 (17·78 - 25·11) |
| White-Mixed-Other | 1390 | 9·28 (8·65 - 9·95) | 1300 | 10·36 (9·68 - 11·09) |
| **Relationship status** | | | | |
| Single | 130 | 10·61 (8·77 - 12·79) | 170 | 10·54 (8·95 - 12·37) |
| Married, civil partnership or cohabiting | 940 | 10·03 (9·28 - 10·84) | 1110 | 11·48 (10·63 - 12·39) |
| Separated, divorced, widowed, or surviving partner | 520 | 11·46 (10·29 - 12·74) | 230 | 11·68 (10·16 - 13·41) |
| **Region** | | | | |
| North East | 80 | 9·74 (8·31 - 11·37) | 80 | 12·02 (9·99 - 14·39) |
| North West | 190 | 9·61 (8·34 - 11·05) | 180 | 10·46 (9·05 - 12·06) |
| Yorkshire and the Humber | 150 | 9·92 (8·50 - 11·55) | 140 | 11·40 (9·46 - 13·69) |
| East Midlands | 160 | 11·71 (9·98 - 13·70) | 150 | 13·42 (11·34 - 15·82) |
| West Midlands | 220 | 12·79 (11·03 - 14·78) | 210 | 14·17 (12·16 - 16·45) |
| East of England | 170 | 9·60 (8·27 - 11·11) | 170 | 10·93 (9·25 - 12·86) |
| London | 210 | 11·80 (10·01 - 13·87) | 170 | 10·46 (8·55 - 12·72) |
| South East | 240 | 9·25 (8·12 - 10·51) | 250 | 11·16 (9·85 - 12·62) |
| South West | 170 | 9·58 (8·20 - 11·15) | 140 | 9·43 (7·98 - 11·12) |
| **Rural-urban classification** | | | | |
| Rural | 340 | 9·80 (8·77 - 10·93) | 300 | 10·15 (9·01 - 11·42) |
| Urban | 1250 | 10·57 (9·83 - 11·35) | 1200 | 11·64 (10·83 - 12·50) |
| **Educational qualifications** | | | | |
| Degree or equivalent qualification | 270 | 8·51 (7·56 - 9·57) | 310 | 8·79 (7·81 - 9·88) |
| Below degree qualification | 770 | 10·57 (9·71 - 11·49) | 750 | 11·69 (10·78 - 12·68) |
| No qualification | 540 | 12·47 (11·17 - 13·91) | 430 | 14·26 (12·61 - 16·07) |

| | | | | |
|---|---|---|---|---|
| **National Statistics Socio-economic Classification (NS-SEC)** | | | | |
| Managerial and professional occupations | 410 | 8·86 (8·02 - 9·78) | 490 | 9·09 (8·22 - 10·05) |
| Intermediate occupations | 430 | 9·57 (8·64 - 10·58) | 330 | 11·53 (10·23 - 12·98) |
| Routine and manual occupations | 670 | 11·94 (10·90 - 13·05) | 650 | 13·41 (12·24 - 14·67) |
| Other | 50 | 14·43 (9·97 - 20·43) | 20 | 18·23 (8·69 - 34·32) |
| **Self-reported general health** | | | | |
| Very good or good | 990 | 9·69 (9·02 - 10·41) | 1010 | 10·76 (9·99 - 11·58) |
| Fair | 420 | 12·91 (11·50 - 14·47) | 340 | 13·12 (11·68 - 14·70) |
| Bad or very bad | 170 | 11·00 (9·23 - 13·05) | 150 | 12·74 (10·47 - 15·42) |
| **Antidepressants prescribed** | | | | |
| Antidepressants prescribed | 280 | 11·47 (10·10 - 13·00) | 150 | 13·96 (11·81 - 16·44) |
| No antidepressants prescribed | 1300 | 10·22 (9·53 - 10·95) | 1350 | 11·12 (10·37 - 11·92) |
| **Body Mass Index (BMI)** | | | | |
| Not overweight or obese | 380 | 7·77 (7·06 - 8·56) | 280 | 8·74 (7·67 - 9·94) |
| Overweight | 470 | 9·64 (8·74 - 10·63) | 590 | 10·58 (9·61 - 11·62) |
| Obese | 570 | 14·61 (13·16 - 16·19) | 510 | 14·86 (13·40 - 16·44) |
| **Waist circumference** | | | | |
| Very high | 1020 | 13·07 (12·01 - 14·21) | 730 | 13·84 (12·63 - 15·16) |
| Other** | 510 | 7·85 (7·21 - 8·54) | 730 | 9·70 (8·93 - 10·54) |
| **Smoking status** | | | | |
| Current cigarette smoker | 270 | 13·73 (12·19 - 15·43) | 340 | 16·89 (15·09 - 18·85) |
| Ex-regular cigarette smoker | 400 | 8·97 (8·03 - 10·01) | 540 | 11·10 (10·01 - 12·30) |
| Never regular cigarette smoker | 920 | 10·21 (9·40 - 11·07) | 620 | 9·61 (8·75 - 10·55) |
| **Frequency of alcohol consumption** | | | | |
| At least once per week | 600 | 8·17 (7·50 - 8·90) | 820 | 9·74 (9·00 - 10·54) |
| Once or twice per month | 200 | 10·18 (8·95 - 11·56) | 180 | 11·66 (10·03 - 13·52) |
| Infrequently | 370 | 12·07 (10·78 - 13·49) | 240 | 15·03 (12·97 - 17·35) |
| Non-drinker/Not at all in last 12 months | 420 | 14·89 (13·20 - 16·76) | 250 | 15·68 (13·52 - 18·12) |

All estimates are weighted, and all are age-adjusted except where the predictor is age or sex or where there is no predictor (i.e., total population estimates). Weighted sample sizes are rounded to the nearest 10.

* 16-44 years has been aggregated to enable comparability with our estimates for undiagnosed diabetes.

** The Health Survey for England (HSE) uses thresholds to categorise waist circumference, with measurements above 102cm for males and 88cm for females considered very high.

Figure S4. Adjusted odds ratios for non-diabetic hyperglycaemia, among all females, by characteristics

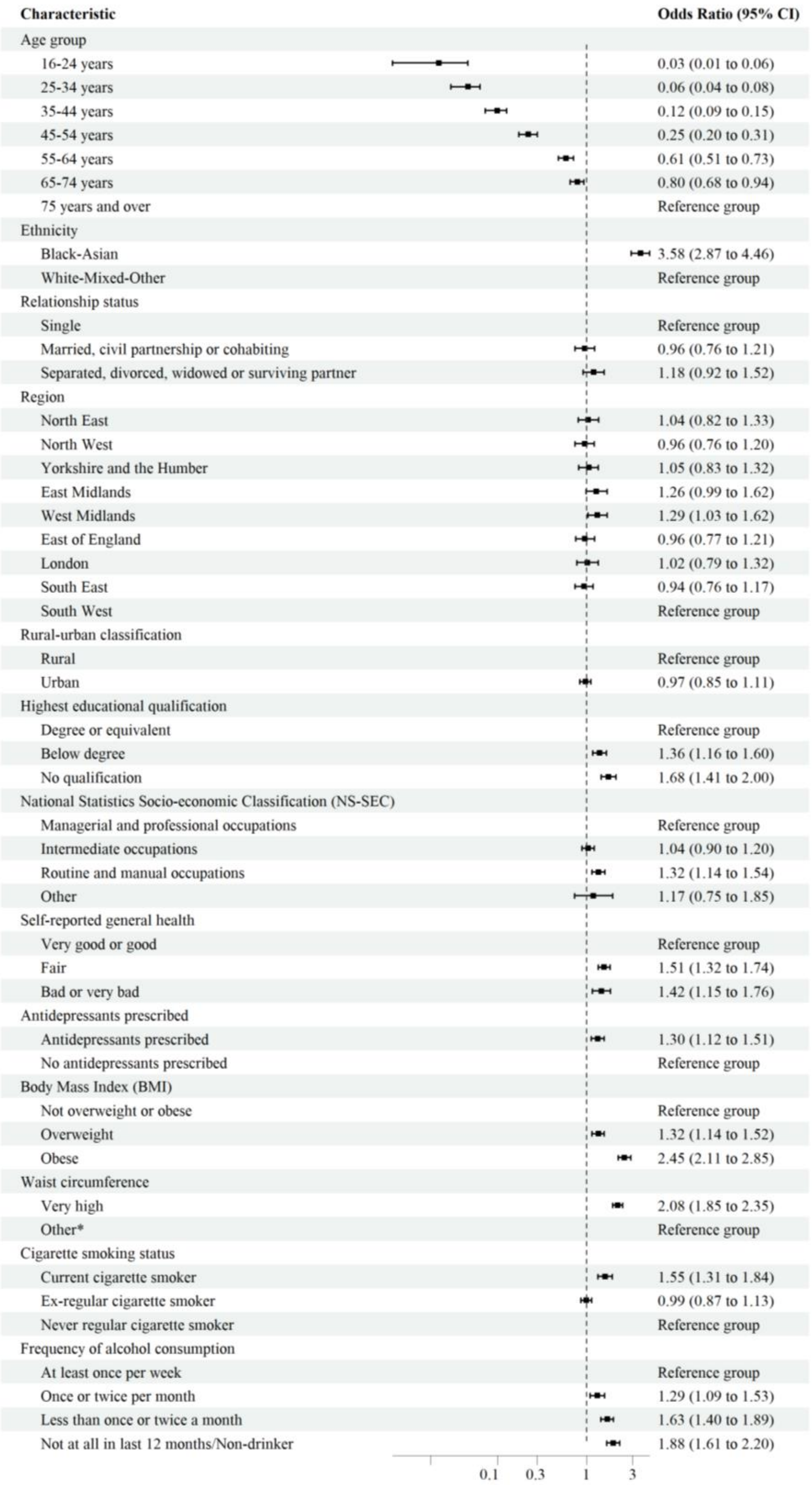


Models were adjusted for age, ethnicity, relationship status, region, rural-urban classification, and highest educational qualification.

* The Health Survey for England (HSE) uses thresholds to categorise waist circumference, with measurements above 102cm for males and 88cm for females considered very high.

Figure S5. Adjusted odds ratios for non-diabetic hyperglycaemia, among all males, by characteristics

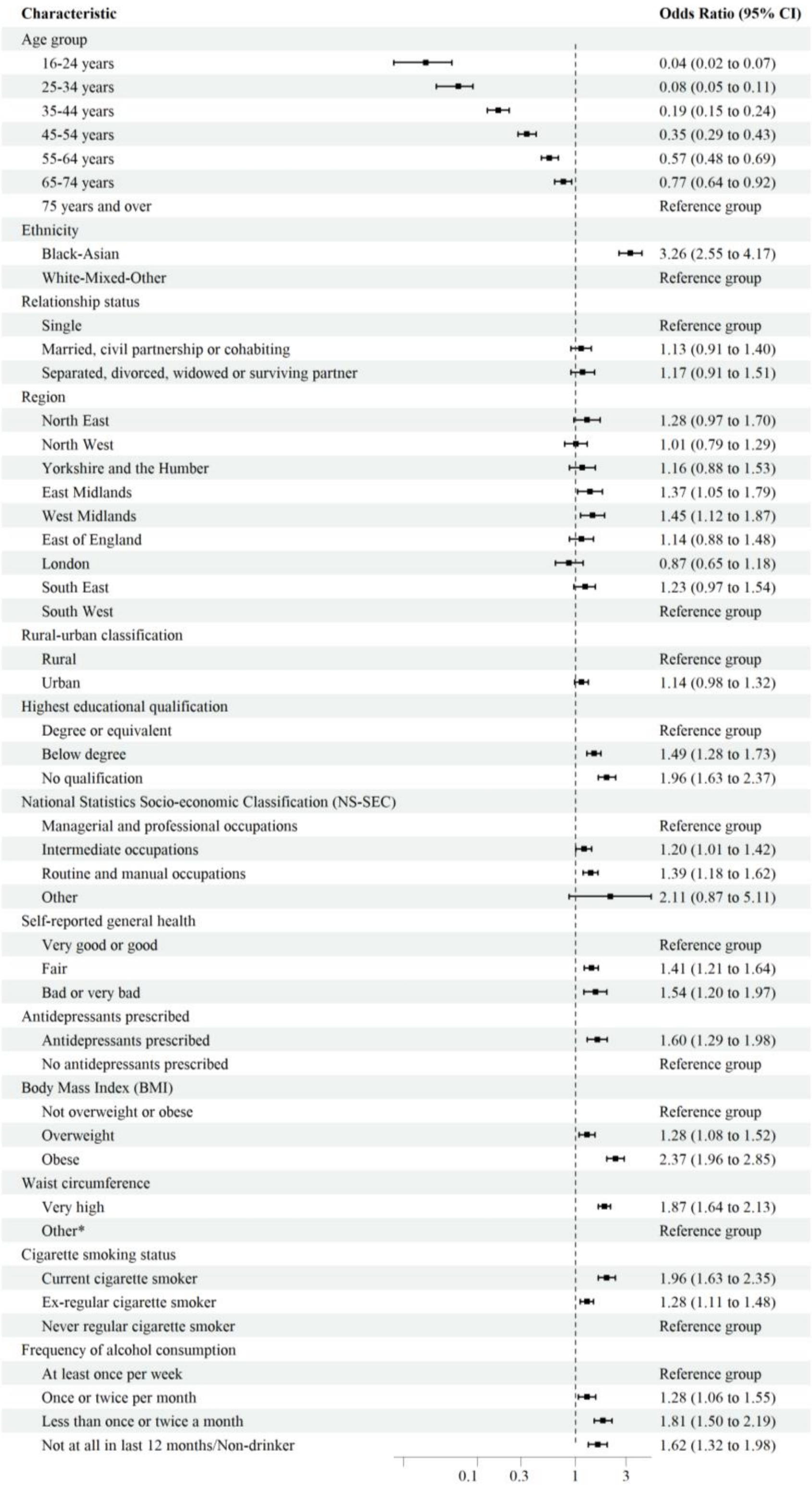


Models were adjusted for age, ethnicity, relationship status, region, rural-urban classification, and highest educational qualification.

* The Health Survey for England (HSE) uses thresholds to categorise waist circumference, with measurements above 102cm for males and 88cm for females considered very high.
